# Prediction of immunotherapy response using live tumor fragments from routine clinical biopsies

**DOI:** 10.64898/2026.06.05.26354635

**Authors:** David Braun, Nicholas Dana, Hilary R. Hernan, Shalini Sahni, Christina Scribano, Christin Johnson, Lindsey Vedder, Erika von Euw, Julie Zweng, Ellen Wargowski, Aishwarya Sunil, Deepa Sharma, Josh Routh, Katherine Rexroad, Payton McDonnell, Victor Jergens, Catarina Costa, Richard Zuniga, Giuseppe V. Toia, Paras M. Patel, Robert C.G. Martin, Umair Majeed, Debabrata Mukhopadhyay, Yanyan Lou, Nima Kokabi, James W. Jakub, David Hays, Andrew K. Godwin, Victoria Giffi, Alexander Gelbard, Andreas Friedl, Emma Kate Duimstra, Roxana S. Dronca, Ruqin Chen, Heather Chalfin, Barbara Broome, Hani M. Babiker, Tarun Chandra, Sean Caenepeel, Laura C. F. Hrycyniak, Chetan Sood, Hilario Ramos, Premal Patel, Pooja Advani, Hinco J. Gierman, Janis Taube

## Abstract

Functional ex vivo assays using live tumor tissues have demonstrated strong predictive accuracy for response to immune checkpoint inhibitors (ICIs) but are not scalable, requiring manual processing of large resections collected at academic centers. Here, an ex vivo live tumor fragment (LTF) platform was developed using standard-of-care biopsies from 228 patients with suspected malignancy collected across prospective, multicenter observational trials and biobanks. Hierarchical clustering of ICI-mediated changes in cytokine production identified two groups: responders and nonresponders. A binary classifier (elive index) using 8 cytokines achieved an AUC of 0.99 for cluster prediction. elive index correctly predicted clinical benefit in 93% (26/28) of patients (*P* = 3.2x10^-5^) and accurately identified 83% (10/12) of objective responders. Critically, elive responders were identified among biomarker-negative patients, highlighting the platform as a scalable approach that complements existing companion diagnostics and expands the population of patients identified to benefit from ICI therapy.

## Introduction

Immune checkpoint inhibitors (ICIs), such as antibodies targeting the PD-1/PD-L1 axis, have become standard therapy for advanced solid tumors and have led to durable responses, with long-term follow-up demonstrating that more than 50% of patients achieve sustained survival in some settings^1,2^. However, current predictive biomarkers lack the sensitivity needed to reliably identify patients who will benefit from treatment, resulting in missed opportunities for these^3^. In parallel, limited specificity contributes to unnecessary toxicity, significant monetary costs and delay in treatment that may provide benefit among nonresponders. Identifying an accurate and clinically implementable predictive biomarker for ICI response is therefore critical to optimizing patient outcomes, minimizing toxicity, and reducing the economic burden of treatment failure.^4^

Current FDA-approved biomarkers for guiding ICI treatment, including PD-L1 expression, tumor mutational burden (TMB), DNA mismatch repair (MMR), and microsatellite instability (MSI), show limited predictive accuracy. Alternative biomarkers such as combination biomarkers (PD-L1 + TMB or TILs), comprehensive genomic tools such as whole exome sequencing, T-cell activation markers, and peripheral biomarkers such as lactate dehydrogenase, have yielded only incremental improvements in predictive accuracy^5–11^. A fundamental constraint of these biomarkers is that they only provide a static, pretreatment snapshot and therefore cannot capture the dynamic functional response of the tumor microenvironment (TME) to therapy^12–14^. Furthermore, intratumoral heterogeneity of PD-L1 expression has been observed in a variety of solid tumor types, including non-small cell lung cancer (NSCLC)^15^ and head and neck squamous cell carcinoma (HNSCC),^16^ leading to discrepancies sufficient to shift PD-L1 scores across clinical guidance cutoffs for ICI therapy.^15^ Cytokines, which serve as key communication signals among immune cells, may offer unique insight into ICI response by capturing the composition and functional status of the TME in individual biopsy specimens. Analyzing these cytokine signals provides a window into tumor immunogenicity, helping to identify patients who are more likely to respond to ICIs^6,17^. The ability to assess real-time changes in the TME may also be critical for improving the accuracy of ICI-response prediction^18,19^.

Ex vivo functional assessment of live tumor tissues, in which the TME architecture and cellular composition are preserved, offers a unique approach by directly measuring how a patient’s tumor responds to ICI exposure rather than inferring response capacity from static features^17,20–24^. Voabil et al. demonstrated that ex vivo cytokine profiling of live, patient-derived tumor fragments (PDTFs) treated with αPD-1 therapy can accurately predict clinical responses in patients. However, these methods require substantial amounts of tissue from surgical resections to address challenges associated with tumor heterogeneity^17^. In addition, the approach is difficult to scale because it requires highly technical expertise and specialized academic centers, limiting its broader accessibility. Expanding this strategy to accommodate smaller tissue specimens, such as core needle biopsies (CNBs) obtained during standard-of-care (SOC) diagnostic procedures, would enable assessment of ICI response in patients with advanced solid tumors, the patient population most likely to receive ICI therapy. Furthermore, developing an assay that is compatible with standard clinical workflows and can be performed in a more automated and scalable manner could substantially increase its clinical utility and impact on patient outcomes.

To address challenges posed by tumor heterogeneity in ex vivo assessment of ICI response in tumor tissues, Ramasubramanian et al. developed an ex vivo tumor profiling platform that preserves the native TME in live tumor fragments (LTFs) cut from CNBs, enabling the functional assessment of immune response to ICI treatment^25^. The platform incorporates a sequential treatment strategy in which ICI-induced cytokine changes to be compared to an IgG control baseline within the same well, thereby reducing the effect of intra-biopsy tumor heterogeneity on measured ICI response. Here, an algorithm was developed (elive index) to predict patient response to ICI treatment, and the correlation of the index performance with clinical outcomes was assessed.

## Results

### Patient study enrollment and specimen collection

To develop a broad classifier for predicting response to immunotherapy, enrollment focused on patients with solid tumors for which ICIs are used clinically in metastatic, recurrent, or neoadjuvant settings (Figure S1A). Fresh tumor tissue was collected via SOC core needle or forceps biopsies from 228 patients enrolled across 3 observational clinical trials (Figure S1B, ClinicalTrials.gov IDs: NCT05478538 [n=16], NCT05520099 [n=48], and NCT06349642 [n=19]) or obtained from tissue biobanks (n = 145). Because enrollment was based on suspected diagnoses, some patients with early-stage disease or tumor types not originally targeted for inclusion were collected and retained in the training dataset to support broad classifier development. Data considered for the data lock were received on or before March 24, 2025.

### Specimen quality and eligibility

At the time of the data lock, 96% of specimens were received in a condition assessable on the platform, with very few samples found to have handling issues (Figure 1, n = 8 [3.7%] or insufficient tissue n = 1, [0.46%]). Because specimen collection occurred prior to confirmation of the patient’s diagnosis, some specimens were excluded from analysis if they were determined to be clinically benign (n = 41 [19%]) or a non-solid tumor type (n = 3 [1.4%]).

**Figure 1.**
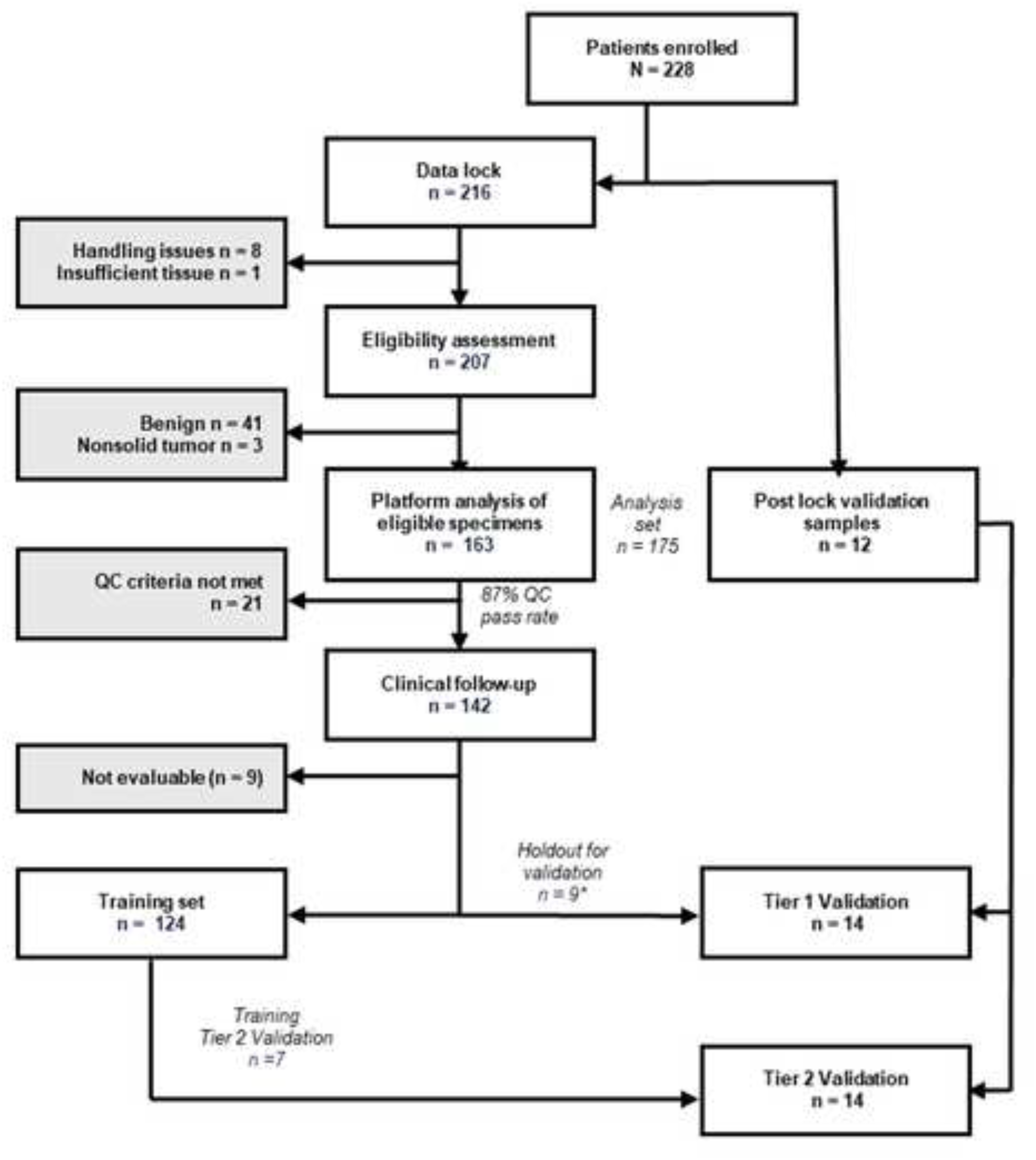
Clinical study flow chart of inclusion and exclusion criteria. Patients were enrolled based on suspected diagnosis and anticipated SOC ICI treatment, with enrollment usually occurring at diagnostic biopsy collection. Of 228 enrolled patients, 216 were included in the elive index development data lock and considered for inclusion in the training dataset, with 12 additional specimens added post–data lock to the validation dataset. Sequential exclusions during eligibility assessment resulted in 163 specimens eligible for platform analysis and a total of 175 specimens included in the study analysis set. Subsequent quality control yielded 142 patients with clinical follow-up and an 87% QC pass rate. Additional specimens were removed for the following reasons: a holdout set of samples for potential validation with clinical results not yet evaluable (n = 5), being QC failures yet yielding a response result after evaluation with the elive index (n = 3), or being sacrificed for targeted QC fail metric development (n = 1). The Tier 1 Validation dataset included patients whose in vivo treatment consisted of immunotherapy that fully matched the ex vivo treatment, as well as patients with a partial treatment match who experienced disease progression. The Tier 2 Validation dataset included specimens with a predicted ex vivo response to ICI in patients who received ICI in combination with another therapy and had a clinical response. The Tier 1 and Tier 2 Validation datasets were used to test the association of elive index with clinical outcome. (N = 28). *One specimen included in the holdout set was determined to be Tier 2 Validation after additional clinical follow-up.

All patients who withdrew consent after the acquisition of the sample (n = 2 [<0.9%]) were also excluded. Of the remaining 163 patients processed on the platform, 142 (87%) met quality control criteria (Figure S2, see methods) for subsequent analysis.

### Training and Validation cohort assignment

Clinical follow-up included both in vivo treatment and outcome data. Because of the observational design of the study, treatment selection was not controlled, and many patients received the ICI evaluated on the elive platform in combination with other therapies (eg, chemotherapy, additional ICIs, or targeted agents). These combinations may contribute to clinical outcomes and were therefore considered when assessing the predictive performance of the elive platform. To account for this, samples were stratified into two validation cohorts. The Tier 1 Validation cohort (n = 14) included 1. patients whose in vivo immunotherapy regimen fully matched the ex vivo condition and 2. patients with partially matched regimens who experienced disease progression. Patients with fully matched regimens were included because their ex vivo response can be directly compared to clinical outcome. Patients with partially matched regimens who progressed were also included because disease progression indicates that the additional in vivo treatments not assessed on the platform failed to provide clinical benefit, thus the ex vivo response to ICI can still be reliably associated with clinical outcome. The Tier 2 Validation cohort (n = 14) comprised patients predicted to respond to ICI monotherapy ex vivo but who received ICI in combination with other therapies in vivo.

A total of 9 patient specimens with an expected in vivo-ex vivo treatment match were withheld from model training in anticipation of inclusion in the Tier 1 Validation cohort with one of these patients reclassified to a Tier 2 Validation (Figures 1). The remaining 124 patients were used for algorithm training and development of the elive index. Following algorithm development, an additional 12 patients were included in the Validation cohort according to study design. Together, the Tier 1 Validation (n = 14) and Tier 2 Validation (n = 14) cohorts (total n = 28) were used to evaluate the association between the elive platform and clinical outcomes.

### Diverse solid tumor types were profiled to develop a broadly applicable ICI response algorithm

To develop a widely applicable response prediction algorithm, we assessed ex vivo responses from a panel of ICI-relevant tumor types. NSCLC was the most common tumor type assessed, making up 40.0% (n = 70) of the total patient population (Table 1). We surveyed patients from diverse demographics such as geographic region, race, ethnicity, age, and sex. The biopsies profiled came from primary lesions (n= 99 [56.6%]), lymph nodes (n = 22 [12.6%]) and distant metastatic sites (n=54 [30.9%]). Ex vivo platform assessments were performed on CNB (n = 117 [66.9%]) or forceps biopsy (n = 58 [33.1%]) samples (Table S1). Whereas approximately half of the CNB specimens arrived with a single core (range 1-4+), two-thirds of the forceps biopsy specimens included 3 forceps bites. The majority of the CNBs were obtained using an 18 G needle (n = 83 [70.9%]), however, a wide range of CNB gauges were successfully processed on the platform (9-22 G). The forceps biopsies processed were collected from mostly standard cup sizes (n = 56 [97%]), however small and large sizes were also processed.

**Table 1.** Analysis set patient characteristics. Table describes enrolled patient demographics, tumor type, primary/metastatic status of the biopsy site, ICI biomarker status, clinical stage at diagnosis, patient treatment and patient treatment response. Other tumor types include peritoneal, thyroid, cervical, breast other than TNBC, lung other than NSCLC, small intestine, neuroendocrine, and esophageal cancers.

| Patient demographics (N = 175) |  |  | Clinical stage at diagnosis |  |  |
| --- | --- | --- | --- | --- | --- |
|  | N | % |  | N | % |
| **Sex at birth** |  |  | I | 33 | 18.9 |
| Male | 94 | 53.7 | II | 4 | 2.3 |
| Female | 80 | 45.7 | III | 24 | 13.7 |
| Unknown | 1 | 0.6 | IV | 83 | 47.4 |
| **Age at diagnosis (years)** |  |  | Unknown | 31 | 17.7 |
| Range | 31-81+ |  | **Patient treatment** |  |  |
| 31-64 | 53 | 30.3 | Monotherapy ICI | 10 | 5.7% |
| 65-80 | 102 | 58.3 | ICI + ICI | 8 | 4.6% |
| 81+ | 18 | 10.3 | ICI + non ICI | 46 | 26.3% |
| Unknown | 2 | 1.1 | Non ICI | 89 | 50.9% |
| **Race** |  |  | Unknown/NA | 22 | 12.6% |
| White | 151 | 86.3 | **Tier 1 Validation + Tier 2 Validation patient treatment response (N = 28)** |  |  |
| Black | 12 | 6.9 | CR, pCR, PR | 12 | 42.9 |
| Asian | 1 | 0.6 | SD | 7 | 25.0 |
| Other/unknown | 11 | 6.3 | PD | 9 | 32.1 |
| **Ethnicity** |  |  | **Tumor type** |  |  |
| Hispanic | 14 | 8.0 | NSCLC | 70 | 40.0 |
| Non-Hispanic | 154 | 88.0 | Kidney | 22 | 12.6 |
| Not reported/unknown | 7 | 4.0 | Head & Neck | 20 | 11.4 |
| **US state of treatment** |  |  | Bladder | 9 | 5.1 |
| Florida | 89 | 50.9 | Skin | 11 | 6.3 |
| Wisconsin | 35 | 20.0 | TNBC | 8 | 4.6 |
| North Carolina | 20 | 11.4 | Hepatobiliary | 6 | 3.4 |
| Arkansas | 10 | 5.7 | Colorectal | 3 | 1.7 |
| Tennessee | 6 | 3.4 | Endometrial | 3 | 1.7 |
| Texas | 2 | 1.1 | Other/unknown | 23 | 13.1 |
| Maryland | 4 | 2.3 | **Biopsy site** |  |  |
| Kansas | 4 | 2.3 | Primary | 99 | 56.6 |
| Kentucky | 2 | 1.1 | Distant metastatic | 54 | 30.9 |
| California | 1 | 0.6 | Lymph node | 22 | 12.6 |
| New York | 1 | 0.6 | **ICI biomarker status** |  |  |
| Oklahoma | 1 | 0.6 | Positive | 40 | 22.9 |
|  |  |  | Negative | 57 | 32.6 |
|  |  |  | Unknown/not applicable | 78 | 44.6 |

### Unsupervised hierarchical clustering identifies two primary clusters of cytokine response to ICI treatment

Voabil et al. previously reported an approach using unsupervised hierarchical clustering of ex vivo cytokine profiling data that revealed two primary sample clusters of cytokine response to ex vivo αPD-1 treatment^25^. One cluster exhibited increased upregulation of cytokines and chemokines, which was subsequently shown to be associated with patient clinical response. Using a similar approach, unsupervised hierarchical clustering was applied to ICI-induced changes in cytokine production rates across the 124 patient specimens (186 total samples) in the training dataset, revealing two primary clusters (Figure 2). The smaller cluster (n = 38 samples [20.4%]; 31 patient specimens [25.0%]) comprised samples with increased analyte parameter values indicative of treatment-induced immunological activity, exemplified by strong induction of IFN-γ and CXCL10. This group was therefore designated as the response cluster. The nonresponse cluster exhibited distinct characteristics such as a response-proximal subgroup with increased VEGF and CCL20 but not IFN-γ or CXCL10 (Figure S3) and a cluster predominantly of head and neck squamous cell carcinoma specimens exhibiting tumor-specific signatures, including upregulation of GM-CSF^26^.

**Figure 2.**
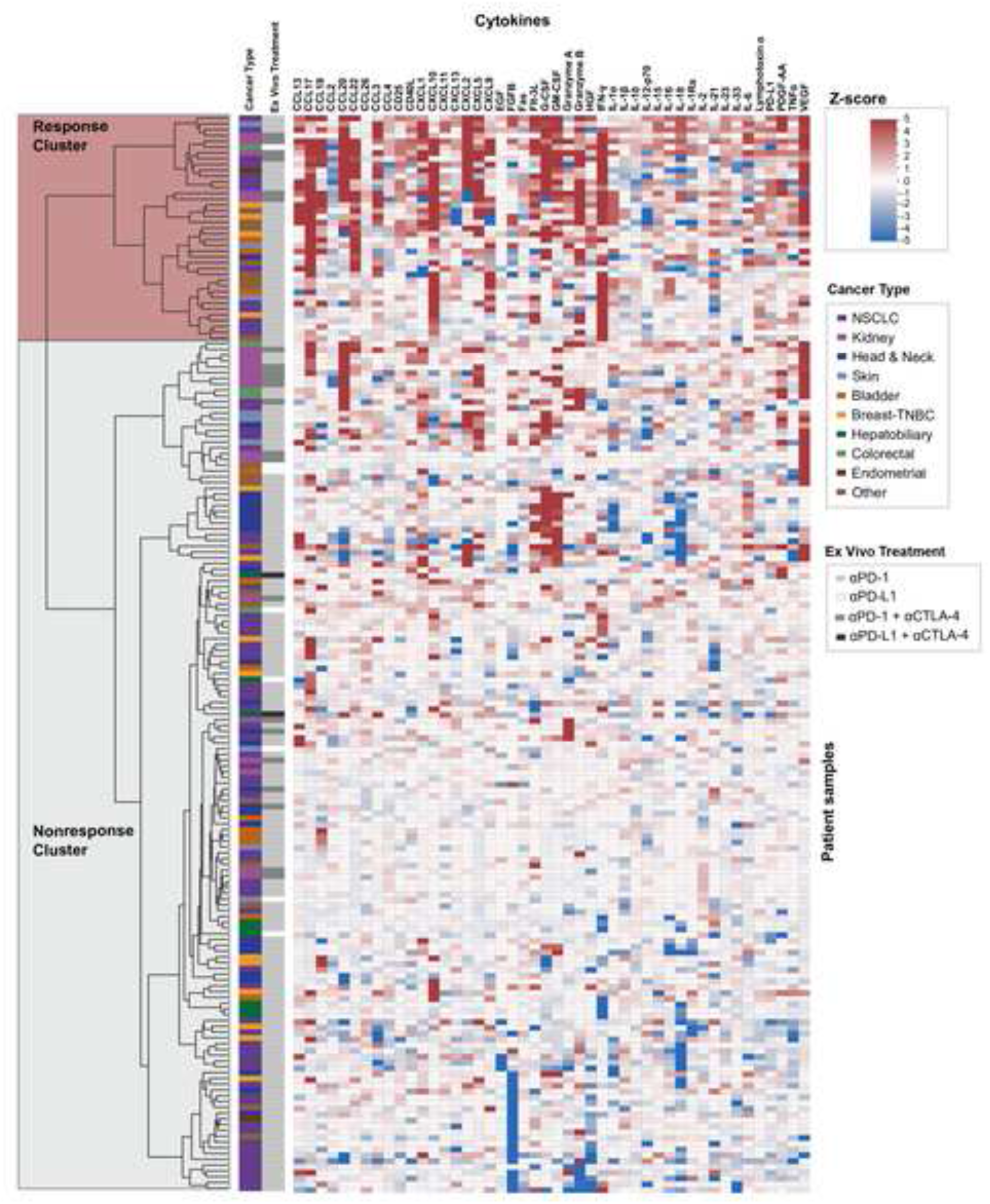
Unsupervised hierarchical clustering of ex vivo cytokine responses to ICI treatment. Heatmap shows z-scores of cytokine measurements across patient samples (n=186 samples from 124 patient tumors) following ex vivo ICI treatment, with hierarchical clustering identifying a response cluster comprising 25% of patients (n= 38 samples from 31 patient tumors) and a nonresponse cluster (n= 148 samples from 93 patient tumors). Heatmap color reflects modified z-scores of the delta in cytokine production rates between the ICI and IgG treatment phases. Annotation bars indicate cancer type and ex vivo treatment.

### Cytokine subcluster analysis reveals 4 distinct immune phenotypes within the response cluster

The response cluster contained 26 analytes significantly upregulated relative to nonresponse samples (Bonferroni-adjusted *P* < 0.05, Table S2), including canonical markers of immune activation such as IFN-γ, CXCL9, CXCL10, TNF-α, and Granzyme B. Hierarchical clustering further identified four distinct subclusters within the response cluster, each characterized by unique cytokine signatures and putative TME states (Figure 3A). Of the 6 specimens with multiple replicates within the response cluster, 5 had replicates that also fell within the same cluster, supporting that assessment across replicate wells yields similar cytokine profiles of response (Figure 3B).

**Figure 3.**
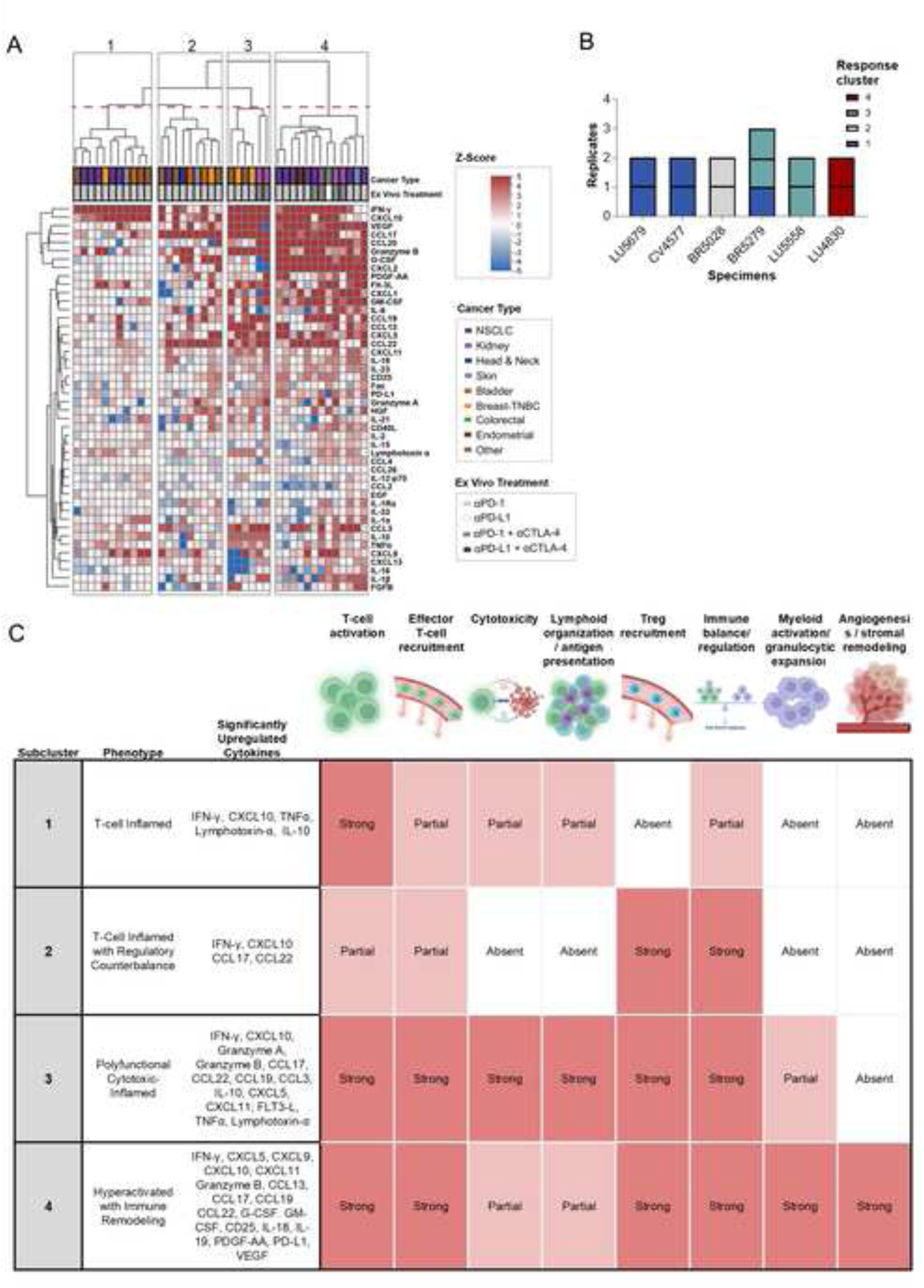
Subclusters within the cytokine response cluster reveal distinct immune response phenotypes associated with ICI-treatment. (A) Hierarchical clustering of cytokine profiles identifies four subclusters within the response cluster with distinct patterns of cytokine upregulation. (B) Replicate mapping of cytokine response profiles is presented for samples where more than 1 replicate mapped to the response cluster. Of the 6 specimens with replicates within the response cluster, 5 had response profiles that were mapped to the same subcluster. (C) Summary table of phenotypic features and significantly upregulated cytokines for each subcluster, highlighting differential engagement of immune mechanisms including T-cell activation, effector T-cell recruitment, cytotoxicity, lymphoid organization / antigen presentation, regulatory T-cell recruitment, immune balance / regulation, myeloid activation / granulocytic expansion, angiogenesis / stromal remodeling. Upregulated cytokines are primarily involved in immune activation and cytotoxic inflammation. Cytokine involvement in each immune mechanism is scored qualitatively as “Strong” when multiple upregulated cytokines contribute to that mechanism, “Partial” when one or a limited subset of contributing cytokines is upregulated, and “Absent” when no upregulated cytokines contribute to that mechanism.

Subcluster 1 represented a T-cell inflamed phenotype, defined by increased IFN-γ, CXCL10, TNF-α, IL-10, and lymphotoxin-α, indicative of active T helper type 1 signaling, immune cell recruitment, and coordinated inflammatory responses (Figure 3C). Subcluster 2 exhibited a T-cell inflamed phenotype with regulatory counterbalance, characterized by enrichment of CCL17 and CCL22 alongside the Th1 IFN-γ and CXCL10, suggesting concurrent recruitment of effector and regulatory T-cell populations. Subcluster 3 demonstrated a polyfunctional cytotoxic-inflamed phenotype, marked by broad upregulation of cytokines and chemokines associated with effector function (granzyme A/B, TNF-α), immune recruitment (CXCL10, CXCL11, CCL19), and antigen-presenting cell support (Flt-3 ligand), alongside regulatory signals such as IL-10. This profile consists of a highly activated immune microenvironment with ongoing cytotoxic activity and feedback regulation. Subcluster 4 represented a more robust TME with a hyperactivated phenotype with immune remodeling, combining features of T-cell activation and cytotoxicity (IFN-γ, CXCL9/10, Granzyme B) with elevated myeloid- and stromal-associated factors, including G-CSF, GM-CSF, VEGF, PDGF-AA, IL-18, and IL-23. This pattern suggests an immune-inflamed TME that is partially constrained by myeloid recruitment, angiogenesis, and compensatory resistance mechanisms.

Collectively, these subclusters span a spectrum of immune activation states within the response cluster, ranging from canonical Th1 T-cell inflamed phenotypes to more complex, polyfunctional TMEs with concurrent regulatory and stromal influences. Despite this heterogeneity, all subclusters share core features of IFN-γ–driven activation and cytokine-mediated immune recruitment, supporting their classification as ICI-responsive phenotypes.

### Enrichment of the response cluster for PDTF-defined immunological responders

To confirm that the response cluster contained samples exhibiting immunological responses to ICI treatment, the Voabil et al. PDTF scoring algorithm which was previously shown to predict clinical response to ICI therapy^17^ was re-engineered and adapted for application to elive-generated data (Figure S4A) and used to classify all samples in the training heatmap dataset. elive-generated data were mapped into the Voabil et al data space, enabling application of the re-engineered PDTF scoring framework (Figure S5). Because the T-cell activation marker CD137 and the cytokine IL-17F are not measured on the elive platform, specimens which fell between a PDTF score of 20 and 40 were identified as borderline specimens, since the inclusion of these missing data points could affect the predicted response. PDTF-responders (PDTF-R) were predominantly localized to the heatmap response cluster while the larger nonresponse cluster was depleted of PDTF-R samples (Figures S4B-D and S3). This enrichment remained statistically significant regardless of how borderline specimens were categorized (*P* < 1 × 10⁻^16^, Figures S4B-D), supporting the validity of hierarchical clustering for predicting immune response in biopsy specimens.

To assess the transferability of the Voabil approach to elive platform data, we evaluated the validation set using a modified Voabil classifier (MVC; Figure S6). In this cohort (n = 32), MVC predictions were concordant with clinical response in 21 of 23 cases, including patients treated with both monotherapy and combination ICI regimens. These findings recapitulate the predictive performance reported by Voabil et al. and provide additional support that specimens within the heatmap response cluster are enriched for true clinical responders.

### Identification of discriminative analytes and creation of the elive index

Application of the MVC to predict clinical outcomes using elive-generated data was suboptimal due to differences in cytokine measurement assay sensitivity and the analytes measured. Accordingly, the elive index was developed to be specifically optimized for the elive platform data. To develop the elive index for clinical outcome prediction, the heatmap-response labels were leveraged to identify discriminative analytes and determine feature thresholds and weights (Figure 4A). Discriminatory performance for each cytokine was evaluated using area under the curve (AUC) analysis, including both receiver operating characteristic (ROC) and precision–recall (PRC) curves (Figure S7). Because samples within the heatmap response cluster comprised 20% (n= 38 samples) of the dataset, PRC analysis provided an important complementary measure in the context of class imbalance. Ranking analytes by AUROC (Figure 4B) and AUPRC (Figure 4C) identified a subset with robust discriminatory performance. Using predefined thresholds of 0.7 for AUROC and 0.6 for AUPRC, 8 cytokines met criteria for inclusion as discriminative features in the elive index algorithm: IFN-γ, CCL22, CXCL10, Lymphotoxin-α, TNFα, CCL17, Granzyme B and IL-10 (Table S3). Analytes with strong predictive capacity demonstrated values approaching 1.0 (eg, IFN-γ; Figure 4D, left panels), whereas values near random classification (AUROC= ∼0.50; AUPRC= ∼0.2, reflecting the positive class frequency) indicated minimal discriminatory power (eg, CCL2; Figure 4D, right panels). Enrichment thresholds for each discriminative analyte were then defined using Matthews Correlation Coefficient (MCC) to optimize classification performance (Figures S8A-D). A sample was classified as enriched for a given cytokine when its normalized delta-rate exceeded the corresponding enrichment threshold. Finally, feature weights were assigned based on discriminatory strength where analytes with a heatmap specificity >0.9 and sensitivity >0.5 were assigned double weight (Table S3). Consequently, enrichment for IFN-γ, CCL22, CXCL10, Lymphotoxin α and TNF-α contributes more substantially to the composite elive index than enrichment for CCL17, Granzyme B, or IL-10.

**Figure 4.**
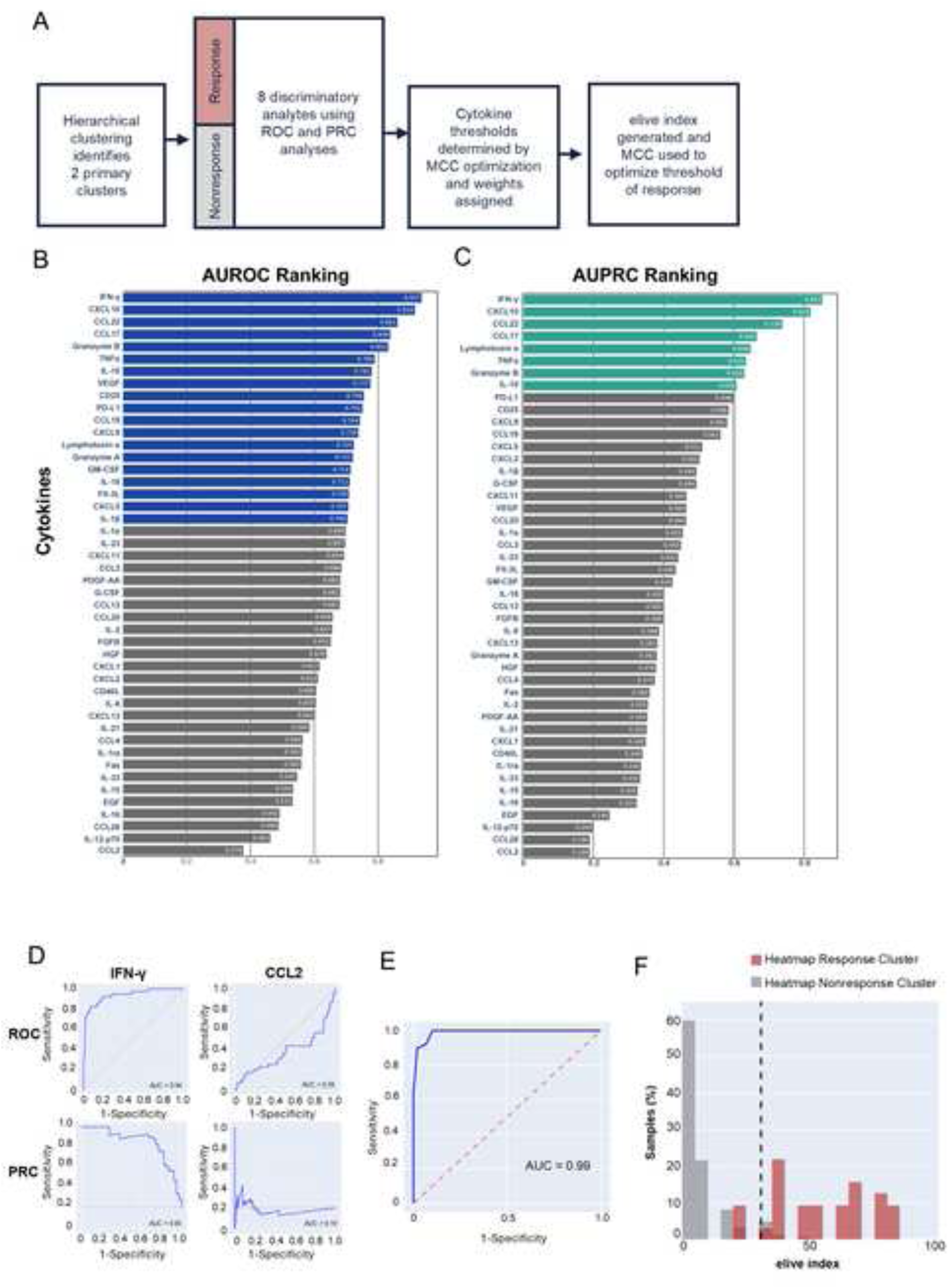
Assessment of the upregulated cytokines in the response cluster reveals a subset that strongly discriminate between the responding and nonresponding clusters of the heatmap. A. Flow diagram illustrating development of the elive index from ICI-induced cytokine response data, including Ward’s hierarchical clustering to define response and nonresponse clusters, identification of 8 discriminative cytokines by ROC and PRC analyses and establishing cytokine thresholds and weights for generation of a composite response index. B. AUROC and C. AUPRC ranks for the 46 analytes assessed. Cytokines falling above the defined cutoffs for ROC and PRC are colored in blue and teal respectively. D. Examples of AUROC (top) and AUPRC curves (bottom) for a cytokine which highly discriminates between the responding (IFN-γ; Left) and nonresponding heatmap clusters and a poorly discriminative cytokine between the responding and nonresponding heatmap clusters (CCL2; Right). E. AUROC showing elive platform performance at predicting the location of the samples within the response and nonresponse clusters of the heatmap (AUC = 0.99). F. Histogram showing the distribution of elive indexes for samples falling within the heatmap response cluster and nonresponse cluster. Dashed black line represents the elive index threshold of response ( > 30.8).

### Composite elive index demonstrates high predictive accuracy of sample location to the response and nonresponse clusters

While some cytokines exhibit exceptional discriminatory power, the elive index algorithm achieves near-perfect performance at predicting the heatmap response label with an AUROC of 0.99 (Figure 4E). This indicates that, while cytokines like IFN-γ may be highly predictive of heatmap cluster, there is at least some complementary information gained by leveraging additional analytes. This is to be expected, as hierarchical clustering utilizes information from all features, even though some may not correlate with position in the larger heatmap. The resulting distribution of scores for the samples within the heatmap response and nonresponse clusters demonstrated high separability (Figure 4F). Evaluation of potential models for separation between predicted sample location in the heatmap response and heatmap nonresponse clusters showed MCC resulted in the best performance, supporting its use for threshold selection in the elive index framework (Figure S9 and Table S4). An optimal score threshold of 30.8 differentiates response from nonresponse cluster labels. Thus, an elive index > 30.8 predicts a response to treatment and ≤ 30.8 nonresponse. Application of the elive index algorithm accurately predicted heatmap class for 179 (96.2%) of samples, achieving a sensitivity of 0.90, specificity of 0.98 for final heatmap response / heatmap nonresponse prediction (Table S4). Technical replicates assessed on the platform demonstrated concordant cluster localization across replicates with 78% of specimens showing concordance of elive index outcomes across all replicates (Figure S10). The distribution of the elive indices and predicted response classifications from training specimens cover the entire range of 0 – 100, with elive platform responder indices being approximately uniformly distributed between 30 – 100 (Figure S11). Use of the full scoring range supports robust classifier performance and discrimination.

### Discordance between standard ICI biomarkers and elive platform outcomes identifies patients with potential to benefit from ICI therapy

Given the limited predictive accuracy of current FDA-approved biomarkers, such as PD-L1 positivity which demonstrates objective response rates of only 20–30% in certain tumor types^23^, we hypothesized that elive platform predictions would meaningfully diverge from standard companion diagnostic testing (ICI biomarkers). To evaluate this, the elive platform response and ICI biomarker status were compared in patients for whom ICI biomarker status was known from clinical records. Among these patients, the elive index identified a substantial proportion of likely responders within both biomarker-positive and biomarker-negative groups (Figure 5A). Comparison across all specimens showed a 44% discordance between ICI biomarker and elive platform response. Notably, 15% (n = 12) of patients were ICI biomarker-negative and classified as responders by the elive platform, suggesting potential missed treatment opportunities using current biomarker strategies alone (Figure 5B). Analysis of treatment patterns further supported this observation as 12.5% of elive platform responders were not treated with ICI (Figure 5C).

**Figure 5.**
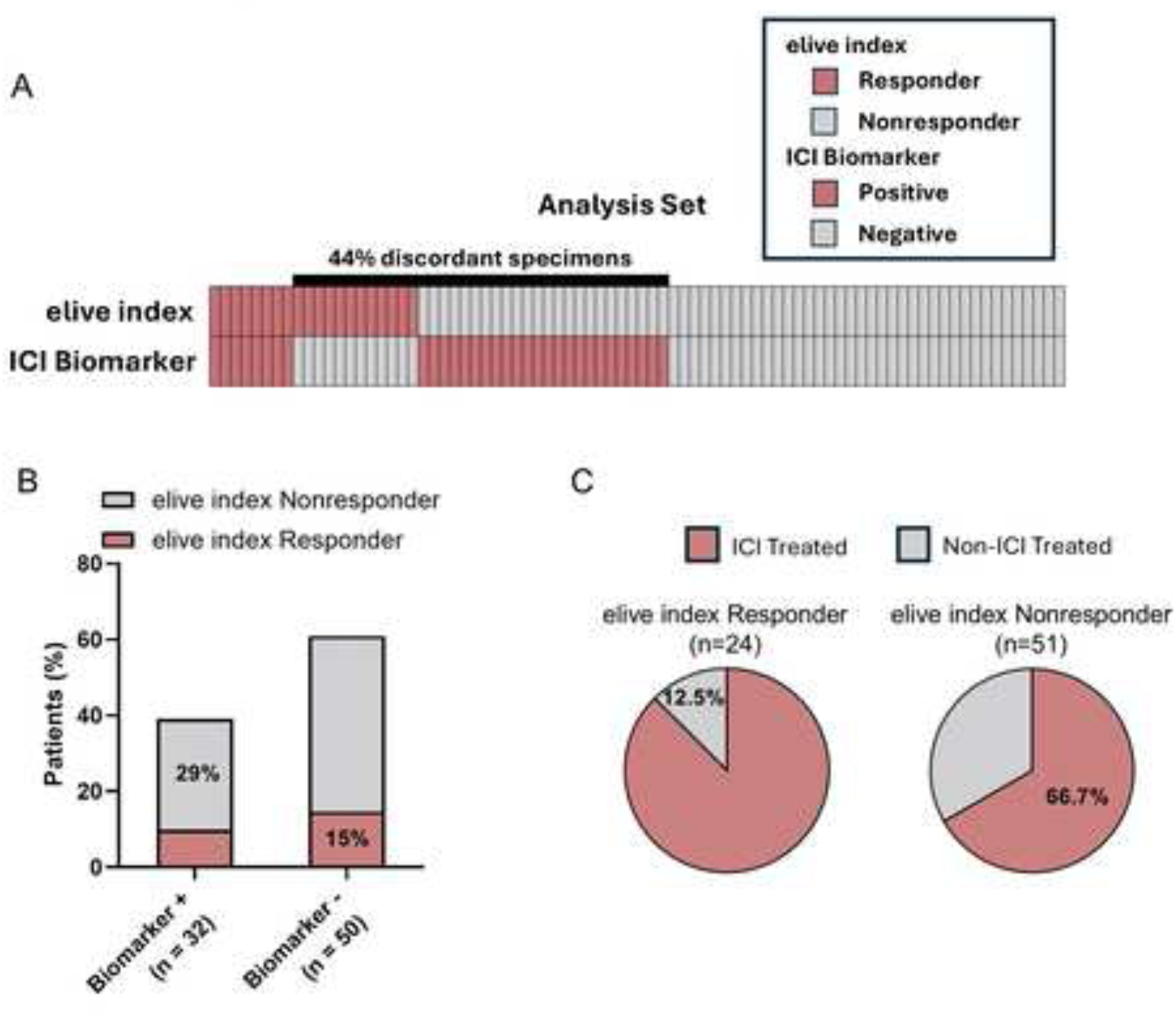
Discordance between companion diagnostic biomarkers and elive index response predictions reveals potential opportunities to improve treatment guidance. (**A**) Box plot of elive index predicted ICI response and ICI biomarker status for all patients with both data points in analysis set. (**B**) Stacked bar plot showing the percentage of patients classified as predicted responders or nonresponders by the elive index stratified by clinical biomarker status (ICI biomarker-positive vs. ICI biomarker-negative) for all evaluable data. (**C**) Distribution of ICI versus non-ICI treated patients among elive index-classified responders and nonresponders for all evaluable data.

### elive platform response accurately predicts patient outcomes in 93% of validation patients

In the validation cohort of 28 patients treated with ICI, the elive platform demonstrated strong predictive performance, correctly classifying 93% (n = 26) of patients (Figure 6A). In the concordance analysis, a weighting scheme was used to discount the contribution of Tier 2 Validation samples. Irrespective of Tier 2 Validation weight (ranging from 0-1) concordance was found to be significant (Weight = 0; Figure S12 *P* = 8.27 x 10^-3^ and Table S5). All 17 (100%) patient specimens with predicted response on the elive platform were collected from patients who achieved disease control with ICI-treatment regimens. This included two patients (KI6173 and LI6422) with matched in vivo and ex vivo ICI treatment who were negative for FDA-approved companion biomarkers (Table S6). Among the elive platform predicted responders, clinical outcomes included 2 complete responses (CR), 3 pathological complete responses (pCR), 5 partial responses (PR), and 7 cases of stable disease (SD) with 3 patients exhibiting SD for greater than 6 months (Figure 6A). In contrast, 82% (9 of 11) elive platform-predicted nonresponders experienced progressive disease (PD), with the majority progressing within the first 4 months of treatment initiation (Figure 6B). This separation in clinical trajectory was reflected in progression-free survival (PFS) analysis for patients receiving ICI only, either monotherapy or in combination, where the median PFS for elive platform-predicted nonresponders was 2.7 months, while median PFS for elive platform-predicted responders was not reached over the follow-up period (Figure 6C). Data in the validation cohorts span 9 tumor types assessed ex vivo with αPD-1/αPD-L1 monotherapy or αPD-1/αPD-L1 plus αCTLA4 combination therapy. QC criteria successfully excluded three specimens that would have produced false-negative predictions; these were drawn from patients with matched ICI treatment who achieved disease control (Figure S13). These findings demonstrate that the elive index robustly predicts clinical benefit from ICI therapy across multiple tumor types and ICI-containing treatment regimens, supporting its translational relevance as a functional response classifier.

**Figure 6.**
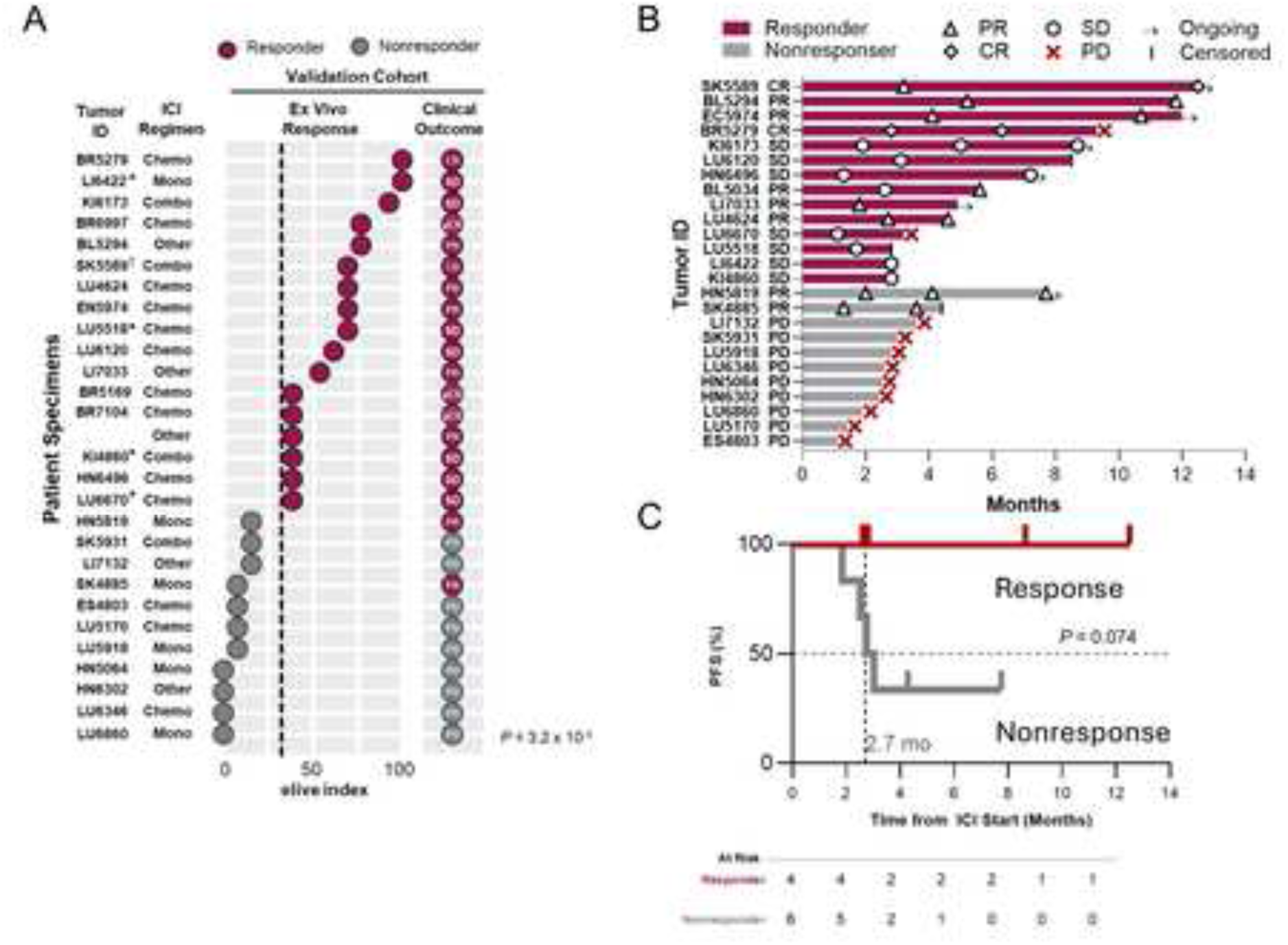
elive index accurately predicts clinical response to ICI treatment in 93% of patients. (A) elive index and accompanying clinical response (best overall response) for all specimens and patients where there was an ICI treatment match between the ex vivo and clinical settings. Specimens are classified into two categories (1) Tier 1 Validation = patients whose in vivo treatment included immunotherapy and fully matched the ex vivo treatment, as well as patients with partial treatment match who experienced disease progression; (2) Tier 2 Validation = specimens with a predicted ex vivo response to monotherapy ICI where the patient received ICI in combination with another therapy. KI4860, SK5931 and KI6173 were treated with αPD-1+αCTLA-4; SK5589 and LI6422 were treated with αPD-L1 and all other specimens were treated with αPD-1 ex vivo. Broken black line represents elive index threshold used to define platform predicted responders (red dots, elive index >30.8) and nonresponders (gray dots, elive index ≤30.8). Clinical outcome is reported as CR = complete response, cPR = complete pathological response, PR = partial response, SD = stable disease and PD = progressive disease. Statistical concordance of the elive index response prediction and clinical outcomes was compared using a weighted binary logistic regression model considering disease control (CR, pCR, PR and SD vs PD, *P* = 3.2 x 10-5). (B) Swimmer plot showing time to progression and clinical course for each patient, stratified by predicted responders and nonresponders. Symbols denote best overall response. (C) Kaplan-Meier analysis of progression-free survival (PFS) from the time of ICI initiation, comparing elive predicted responders with nonresponders with removal of patients with pCR. Median PFS for nonresponders was 2.7 months; median PFS for responders was not reached. BL, bladder; BR, breast; EN, endometrial; ES, esophageal; HN, head and neck; KI, kidney; LI, liver; LU, lung; SK, skin (including cutaneous squamous cell carcinoma and melanoma); OT, other. *<6 months SD; †Tier 2 Validation ICI combinations; all other ICI combinations are Tier 1 Validations.

### Representative patient cases further illustrate the clinical utility of the elive platform in guiding ICI treatment decisions

#### Patient with HNSCC and progressive disease with pembrolizumab treatment

A patient with a history of HPV (p16)-positive squamous cell carcinoma (SCC) of the tonsil developed a new lung lesion confirmed to be metastatic SCC (Figure 7). The tumor was PD-L1 negative (CPS<1). Two 20G CNBs from the lung metastasis collected prior to treatment initiation yielded sufficient tissue for a single ex vivo assessment well, which was treated with αPD-1. Cytokine profiling revealed minimal induction of discriminative analytes; CCL17 was the only response-associated cytokine exceeding the threshold (Figure 7A). Following progression on prior platinum-containing therapy (Figure 7 B), the patient was treated with pembrolizumab monotherapy despite the absence of PD-L1 expression. At the 3-month radiographic assessment, the patient exhibited progressive disease, with increase in tumor burden (Figure 7C). The resulting elive index was 7, below the predefined platform response threshold of 30.8, indicating a predicted elive platform nonresponse. Histological evaluation demonstrated clusters of carcinoma cells consistent with metastatic SCC (Figures 7D; left, middle and S14A) as well as the presence of immune cells within the TME (Figures 7D, right and S14B) supporting an immune-infiltrated, but functionally ICI unresponsive state. This case exemplifies concordance between a low elive index and lack of clinical benefit from ICI, highlighting the potential utility of the platform in identifying patients unlikely to respond to ICI and who may benefit from alternative therapeutic strategies.

**Figure 7.**
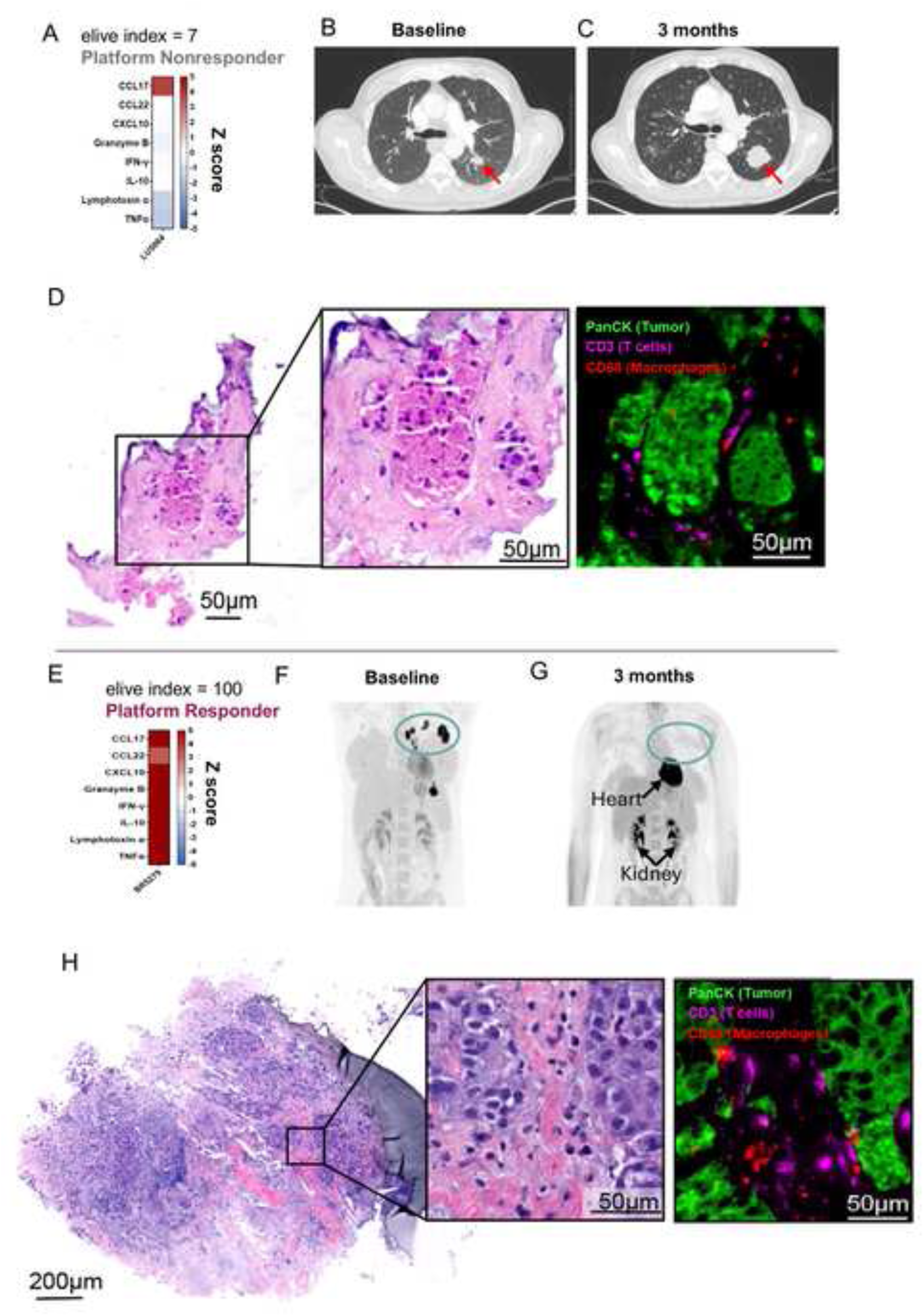
Application of the elive index to ex vivo cytokine response profiles predicts real-world response outcome to ICI therapy. (A) Ex vivo cytokine profiling of a 20G CNB specimen cut, separated into 1 sample replicate and treated with αPD-1 yielded an elive index of 7, below the responder threshold of 30.8 and classifying this patient as a predicted nonresponder. (B) Baseline and (C) 3-month follow-up CT scans from a patient with HNSCC(HN5064) treated with pembrolizumab and chemotherapy. This patient exhibited disease progression by RECIST criteria at 3 months. (D) H&E (left, middle) and mIF (right; with PanCK [tumor, green], CD3 [T cells, magenta], CD68 [macrophages, red labeling of a representative LTF following ex vivo treatment. (E) Cytokine response profiles from three replicate samples following ex vivo treatment with αPD-1 yielded an elive index of 100, classifying this patient as a predicted responder (sample used for elive index outlined in black). Heatmap color reflects Z-scores of the delta in cytokine production rates between the ICI and IgG treatment conditions. (F) Baseline and (G) 3-month follow-up FDG-PET scans from a PD-L1-positive patient with de novo oligometastatic TNBC (BR5279). This patient achieved a CR to pembrolizumab and chemotherapy at 3-month follow-up. Tumor tissue obtained via 4, 14G CNBs prior to treatment initiation was profiled on the platform. Arrows indicate physiologic FDG uptake in heart and kidneys; the previously avid lesions (circled) have resolved (H) H&E (left, middle) and mIF (right; with PanCK [tumor, white], CD3 [T cells, magenta], CD68 [macrophages, green]) labeling of a representative LTF following ex vivo treatment.

#### Patient with triple negative breast cancer (TNBC) shows strong therapeutic response

A patient with oligometastatic stage III TNBC involving the primary tumor and multiple lymph nodes was evaluated (Figure 7). Prior to treatment, 4, 14G CNBs were obtained from an axillary lymph node, yielding sufficient tissue for three replicate wells to be assayed on the elive platform. This specimen achieved the maximum elive index of 100, characterized by robust induction of all discriminative analytes in one replicate and consistent activation of cytotoxic T-cell associated cytokines (CXCL10, IFN-γ, TNF-α, lymphotoxin-α, and Granzyme B) across all replicates (Figure 7E). As a borderline PD-L1–positive case (CPS = 10), the patient received neoadjuvant pembrolizumab in combination with paclitaxel and carboplatin (Figure 7F). At three months, the patient achieved a CR (Figure 7G). As a result, surgery was deferred, and the patient continued pembrolizumab monotherapy, maintaining complete response for over 6 months. Histologic analysis confirmed presence of tumor cells (Figures 7H; left, middle and S14C) and abundant CD3⁺ T-cell infiltration, consistent with the observed cytokine profile (Figures 7H, right and S14D). Although chemotherapy was part of the patient’s regimen, such rapid and complete responses are less common with chemotherapy alone^27^ supporting a meaningful contribution from ICI therapy and concordance with the high elive index.

## Discussion

Current FDA-approved biomarkers for ICIs (PD-L1, TMB, and MSI/MMR) demonstrate poor predictive accuracy. Functional ex vivo profiling platforms using live tumor tissue have shown considerable promise for improving response prediction. However, their clinical scalability has been constrained by the need for large amounts of tissue and labor-intensive manual processing. Here, an ex vivo LTF platform and accompanying binary classifier of response are reported, designed for use with a single routine clinical biopsy (core needle or forceps) following overnight shipment to a centralized laboratory from sites across diverse geographic regions. Importantly, the classifier was built using only ex vivo cytokine induction in LTFs and followed by orthogonal clinical validation. Preliminary performance of the classifier suggests high predictive accuracy for clinical response across multiple ICIs and tumor types. In addition, the platform shows potential to complement existing ICI biomarkers by identifying responders among biomarker negative patients.

A key advancement of this platform is its ability to characterize functional cytokine response to ICI treatment in live tumor tissue while preserving the intact TME, providing biological insights into the underlying mechanisms of ICI response. Unsupervised hierarchical clustering of our cytokine profiling data identified two primary immune response clusters (response vs nonresponse clusters). Within the response cluster, four distinct cytokine phenotypes capture complex and dynamic immune states, including inflamed, cytotoxic and regulatory programs, rather than a single mechanism of action. These phenotypes help to provide key insights into the immune cell content and dynamics in these TMEs elucidating the biological processes involved in the response to ICIs.

Current FDA-approved biomarkers for ICIs provide static snapshots of the highly dynamic TME and fail to capture treatment-induced changes in immune activity, limiting their ability to fully predict therapeutic response. In contrast, the elive platform enables longitudinal assessment of treatment-induced immune activation, thereby addressing a key limitation of existing biomarkers and providing a more functional measure of response potential. The elive index, derived from the 8 most discriminative cytokines for distinguishing between the response and nonresponse clusters, demonstrated high concordance with clinical benefit in a validation cohort of patients. Importantly, the platform also identified discordance with established biomarkers in 44% of training dataset specimens, including 15% of biomarker-negative patients who were predicted as responders, suggesting that functional profiling may complement existing biomarkers and identify patients who may benefit from ICI despite being negative by standard testing.

Unlike other LTF ex vivo profiling approaches, which require large surgical specimens and specialized infrastructure, this method is compatible with routine clinical sampling. The elive platform is readily translatable to clinical workflows, utilizing core needle biopsies and other standard of care specimen collection formats, including forceps biopsies. Although the amount of tissue available from these specimen formats is limited, application of a sequential treatment strategy where control (IgG) and ICI treatments are performed sequentially on the same tissue in a single well addresses challenges associated with intra-specimen heterogeneity. Additionally, projected turnaround times of <7 days between tissue collection and report generation support use of platform results in real-time clinical decision making.

The data reported here provide strong evidence that the elive platform can be run at scale with a high success rate (QC pass rate >87%), processing more than 200 specimens collected from multiple clinical sites ranging in size from major academic medical centers to smaller community-based settings. Forceps biopsies only failed on assay QC, while CNB biopsies failed mostly for specimen QC (Figures S2 E-F).The platform also demonstrates evidence for compatibility across multiple tumor types and checkpoint inhibitor regimens, highlighting the potential use of a universal classifier to predict ICI response. Additionally, the ability to process multiple specimen form factors suggests that the platform could be adapted for use with additional biopsy types such as fine needle aspirates and punch biopsies. The use of a limited cytokine panel in the elive index also points to opportunities for future assay simplification and additional scalability. Lastly, physical deployment of the platform at clinical sites would provide an opportunity to reduce operational turnaround times and has the potential to further improve tissue quality and platform performance.

Although the results reported here are promising, some limitations of this study warrant consideration. First, the elive index was developed using ex vivo molecular response data rather than direct clinical outcomes. Although the cytokine-based classifier demonstrated strong concordance with clinical response across the validation cohort, training on molecular endpoints introduces an inherent assumption that ex vivo immune activation recapitulates clinical benefit. Future iterations of the platform will incorporate clinical outcome data directly into model training, which may further refine the relationship between ex vivo response signatures and durable patient benefit. Second, the validation cohort was heterogeneous with respect to tumor type, ICI regimen, line of therapy, and biopsy collection method. Although this diversity supports the generalizability of the platform across clinical contexts, it also limits the statistical power to evaluate predictive performance within any single tumor indication or treatment setting. Larger, indication-specific cohorts will be needed to characterize the platform’s performance and establish indication-level cutoffs. Third, as an observational study, the analysis is subject to confounds inherent to real-world treatment data. Patients received a range of ICI regimens, often in combination with chemotherapy, targeted agents, or other immunomodulatory therapies, and the contribution of these concurrent treatments to clinical outcome cannot be fully distinguished from the ICI effect that the ex vivo assay was designed to model. Prospective validation in controlled clinical trial settings will be required to fully establish the clinical utility of the elive platform and to support its incorporation into treatment decision-making.

Beyond addressing these limitations, several directions for future development warrant exploration. The current platform was validated primarily against αPD-1, αPD-L1, and αCTLA-4 monotherapy and dual checkpoint inhibition. Extending the platform to additional immunotherapy mechanisms, including novel combination regimens, bispecifics, engineered cell therapies, and emerging checkpoint targets, would broaden its applicability across the rapidly evolving immuno-oncology landscape. Similarly, expansion into tumor indications underrepresented in the current cohort, particularly those with historically low ICI response rates where biomarker-guided patient selection is most needed, would fill critical gaps in the treatment landscape. The elive index itself may also benefit from continued refinement. Incorporation of additional ex vivo features, such as broader cytokine and chemokine panels, single-cell phenotypic readouts, or spatial measures of immune engagement, could capture aspects of the antitumor response not reflected in the current classifier. Likewise, alternative modeling approaches, including ensemble methods and deep learning architectures trained on larger datasets, may yield further gains in predictive accuracy as the platform matures.

Together, these findings demonstrate the feasibility and clinical utility of using a functional ex vivo platform for assessing tumor immune responsiveness using routine biopsy specimens. By capturing dynamic, treatment-induced immune activity within the intact TME, this approach provides biological insight beyond static biomarker strategies. The platform therefore offers a method with the potential to improve patient stratification for ICI therapy, including cases where established biomarkers are absent or inconclusive. Collectively, these results support the advancement of functional precision immuno-oncology as complementary tools to existing biomarker paradigms for guiding immunotherapy treatment decisions.

## Data Availability

All data produced in the present study are available upon reasonable request to the authors.

## Resource availability

Further information and resources are available upon request.

## Acknowledgements

We thank the patients and their families for their participation in this research, and the clinical site investigators, study coordinators, and pathology teams whose contributions made this work possible. We thank Christina Vivelo for editorial assistance and critical review of the methods section, and Mikaela Schultz for laboratory management and supervision of the experimental work.

## Author Contributions

Conceptualization: D.B., N.D., H.R.H., C.J., S.C., L.C.F.H., C.S., H.J.G.

Methodology: N.D., H.R.H., S.S., C.M.S., C.J., L.V., E.vE., J.Z., J.R., C.C., A.F., L.C.F.H., C.S., H.J.G.

Software: N.D., C.J., C.S.

Validation: C.J., A.S., P.M., D.M.

Formal Analysis: N.D., S.S., C.M.S., C.J., L.V., E.vE., J.Z., T.C., S.C., L.C.F.H., C.S.

Investigation: N.D., S.S., C.M.S., C.J., E.W., A.S., K.R., P.M., A.F.

Resources: H.R.H., S.S., C.J., D.S., C.C., R.Z., G.V.T., P.M.P., R.C.G.M., U.M., D.M., Y.L., N.K., J.W.J., D.H., A.K.G., V.G., A.G., A.F., E.K.D., R.S.D., R.C., H.C., B.B., H.M.B., P.A.

Data Curation: N.D., H.R.H., S.S., C.M.S., C.J., L.V., E.vE., J.Z., A.S., D.S., K.R., P.M., V.J., C.C., S.C., L.C.F.H., C.S.

Writing – Original Draft: D.B., N.D., H.R.H., S.S., C.M.S., C.J., L.V., E.vE., J.Z., S.C., L.C.F.H., C.S., H.R., H.J.G.

Writing – Review & Editing: D.B., N.D., H.R.H., S.S., C.M.S., C.J., L.V., E.vE., J.Z., E.W., A.S., D.S., J.R., K.R., P.M., V.J., C.C., G.V.T., P.M.P., R.C.G.M., U.M., N.K., J.W.J., D.H., A.K.G., A.F., E.K.D., R.S.D., H.C., B.B., H.M.B., T.C., S.C., L.C.F.H., C.S., H.R., P.P., P.A., H.J.G., J.T.

Visualization: D.B., N.D., S.S., C.J., L.V., E.vE., J.Z., A.F., S.C., C.S.

Supervision: D.B., H.R.H., C.M.S., C.J., S.C., L.C.F.H., C.S., H.R., P.P., H.J.G., J.T.

Project Administration: H.R.H., C.M.S., C.J., L.V., L.C.F.H., H.R.

All authors reviewed and approved the final manuscript.

D.B. and H.J.G. served as corresponding authors due to their scientific leadership in the development, execution and presentation of these data.

## Declaration of Interests

N.D., H.R.H., S.S., C.M.S., C.J., L.V., E.vE., J.Z., E.W., A.S., D.S., J.R., K.R., P.M., V.J., C.C., S.C., L.C.F.H., C.S., H.R., P.P., and H.J.G. are employees of Elephas Biosciences and hold equity, stock options, or both in the company.

S.C. is an inventor on U.S. Patent Application Nos. 12467918B1, and 11366101, which cover technology related to the elive platform. L.C.F.H. is an inventor on U.S. Patent Application No. 11366101.

A.F. holds equity in Elephas Biosciences.

D.B. reports share options in Elephas; advisory board, consulting or personal fees from Cancer Expert Now, Adnovate Strategies, MDedge, CancerNetwork, Catenion, OncLive, Cello Health BioConsulting, PWW Consulting, Haymarket Medical Network, Aptitude Health, ASCO Post and Harborside, Targeted Oncology, Merck, Pfizer, MedScape, Accolade 2nd MD, DLA Piper, Dechert, AbbVie, Compugen, Link Cell Therapies, Scholar Rock, NeoMorph, Nimbus, Exelixis, AVEO, Eisai, Daiichi Sankyo, Caris, Ottimo Pharma, LG Chem, Calico, and Elephas; and research support from Exelixis and AstraZeneca, outside of the submitted work.

D.B. and J.T. serve on the Scientific Advisory Board of Elephas Biosciences and have received consulting fees.

J.T. has participated on advisory boards for AstraZeneca, Bristol Myers Squibb, Merck, Regeneron, Elephas, Lunaphore, Roche and Moderna and has received research grants from Bristol Myers Squibb and Akoya Biosciences.

E.K.D. and T.C. are employees of EmpiriQA LLC, which received consulting fees from Elephas Biosciences for statistical analysis support related to this work.

R.C. received research funding from Elephas Biosciences.

R.Z. has served on advisory boards for Mirati Therapeutics, Takeda, Bristol Myers Squibb, and Genentech, and has served as a paid speaker and investigator for Chemouthpiece. These relationships are outside the submitted work.

G.V.T., P.M.P., R.C.G.M., U.M., D.M., Y.L., N.K., J.W.J., D.H., A.K.G., V.G., A.G., R.S.D., H.C., B.B., H.M.B., and P.A. declare no competing interests.

Elephas Biosciences holds intellectual property related to the elive platform, including the U.S. Patent Applications listed above. The study was designed, conducted, analyzed, and reported by the authors; no editorial control was exercised by parties outside the author group.

All funding was provided by Elephas Biosciences.

Declaration of generative AI and AI-assisted technologies in the writing process.

During the preparation of this work the authors used Claude in order to review for spelling, grammar and consistency. After using this tool/service, the author(s) reviewed and edited the content as needed and take full responsibility for the content of the published article.

## METHODS

### Patients and specimens

Human biopsy collection was approved by institutional review boards and conducted through clinical trials (ClinicalTrials.gov identifiers: NCT05478538, NCT05520099, NCT06349642,) or biobank collaborations (NCT07327489). Eligible adults (≥18 years) had histologically confirmed or suspected solid tumors with a focus on approved indications for ICI therapy (Fig 1). Exclusion criteria included pregnancy, autoimmune disease contraindicating immunotherapy, or active immunosuppression. Tumor types included: NSCLC, SCLC, kidney, head and neck (HNSCC), bladder, skin, TNBC, colorectal, hepatobiliary, esophageal, cervical, endometrial. Biopsies were obtained with informed consent through institutional review board approved protocols using core-needle or forceps techniques from primary or metastatic sites, stabilized immediately in transport media, and shipped overnight in NanoCool biothermal containers (Peli Biothermal, 2-85225) maintaining −2 °C to 10 °C^25^. Biomarker status was determined using PD-L1 or MMR protein expression, MSI and/or TMB from patient records.

Patients were considered evaluable for clinical correlation analysis if they had measurable disease at ICI initiation, a baseline radiographic or pathological assessment, and at least one post-baseline assessment interpretable for response. For patients enrolled in prospective observational trials NCT05478538 and NCT05520099, tumor response was assessed by site investigators using RECIST 1.1^28^ as the primary criterion, with iRECIST applied in parallel to identify ICI-specific atypical response patterns. Imaging was performed as standard of care, with reassessment every 6 to 8 weeks. In cases of divergence between RECIST 1.1 and iRECIST due to pseudoprogression (Unconfirmed progressive disease followed by stabilization or response), the iRECIST-confirmed downstream category was used to avoid misclassification of ICI-mediated atypical response as progression.

For patients sourced from NCT06349642, NCT07327489 or biobank collaborations, RECIST1.1 assessment was used when available and if not, real-world response was determined from site-based review of the longitudinal clinical documentation (treating physician notes, radiology reports)^29^. For patients enrolled in the neoadjuvant study, response was determined by pathological assessment of the surgical resection specimen per protocol, with pCR defined as absence of residual invasive disease.

### Specimen preparation

CNBs (12–22 gauge) ≥ 3 mm in length were sectioned at a 20° angle using an automated proprietary cutting device to generate 300 µm thick live tumor fragments (LTFs) while CNBs < 3 mm and forceps biopsies were manually cut into LTFs. To mitigate variability due to intraspecimen heterogeneity, LTFs were pooled prior to plating into 24-well culture plates. Each sample well was overlaid with hydrogel (elive^TM^ Gel, Elephas) which was polymerized by UV light for 90 seconds, followed by 500 µL of culture medium containing an isotype-matched IgG control (Table S8).

### Treatments

Ex vivo assessment of ICI response followed the sequential treatment protocol detailed previously^25^. IgG and ICI antibodies were used at a concentration of 50 µg/mL. After 24 hours of IgG treatment, ICI was added based on the therapy most likely to be administered clinically for each patient’s tumor type as follows; for NSCLC, HNSCC, colorectal, endometrial, and TNBC: pembrolizumab biosimilar (αPD-1; BioXcell, #SIM0010); kidney: pembrolizumab biosimilar and ipilimumab biosimilar (αCTLA-4; BioXcell, #SIM0004); Liver, atezolizumab biosimilar (αPD-L1; BioXcell, #SIM0009) (Table S7). Any tumor types other than those listed here received pembrolizumab biosimilar. In some cases, when sufficient tissue was available for replicate wells, additional ex vivo treatments were selected to reflect the most likely clinical treatment options based on current guidelines. Upon experiment completion, each well was fixed in 10% phosphate-buffered formalin for standard histology processing including multiplexed immunofluorescence (mIF) staining (Table S8).

### Cytokine profiling

Human XL Cytokine Luminex Performance Assay Premixed Kits (R&D Systems, FCSTM18B-30 and CUSTOM-LXSA-H-17) were used to assay conditioned media collected from individual culture sample wells at designated time points after IgG (4 hours and 24 hours) / ICI (24 hours) treatment for 46 cytokines. In rare cases where cytokine concentrations exceeded detection limits and were reported as null values, these values were replaced with the largest concentration read on the plate. Measurements below the lower limit of quantitation (LLOQ) were left unchanged and included in analyses. To account for supernatant collection and dilution at each timepoint, cumulative concentrations were calculated from adding the values reported by the Luminex assay at each timepoint to the previous timepoint’s value multiplied by the collection fraction. ICI-induced change in cytokine cumulative concentrations was determined by the delta of slope between ICI and IgG control treatment phases (ICI slope - IgG slope).

### Statistical analyses

Bar graphs display the mean and standard deviation, while violin plots depict medians as solid lines, with dashed lines indicating the first and third quartiles.

#### Adaptation of Voabil et al

The Voabil et al. scoring framework was adapted for use on the elive platform with elive native data by reconstructing the thresholds of discriminative analytes (Optuna 3.6.1 optimization package, Python). A randomized, iterative solver was used to predict enrichment thresholds for all 12 discriminative cytokines from Voabil et al. by minimizing the function 1 – R^2^, where R^2^ is the coefficient of determination calculated by comparing the reconstructed vs. actual scores for the entire cohort. The cumulative distribution transform (CDT) was used to transform novel cytokine values generated on the elive platform to the same data space as the Voabil et al. cohort, where the cytokine-specific CDTs were trained on a cohort of CNBs. A “Borderline” classification band was introduced between the platform responder and platform nonresponder classification bands to account for missing a marker of T-cell activation, CD3+ CD137+ and the cytokine IL-17F, that were not measured on the elive platform. The reconstructed Voabil et al. scoring framework was defined as the Modified Voabil Classifier (MVC).

#### elive index

Agglomerative hierarchical clustering was performed to cluster by sample and analyte using Ward’s method on 90% selectively winsorized delta values. Response or nonresponse cluster labels were assigned to samples falling in the hot or cold heatmap macrocluster, respectively. Cytokines were selected as discriminative features based on their power to differentiate the response and nonresponse clusters using area under the ROC curve (AUROC) and area under the precision-recall curve (AUPRC) analyses with the performance criteria of AUROC > 0.7 and AUPRC > 0.6 [Figures 4D and S7]. For each discriminatory cytokine, an optimal enrichment threshold was determined to achieve high performance using Matthews correlation coefficient (MCC). The most discriminative analytes, defined as those with heatmap specificity > 0.9 (90%) and sensitivity > 0.5 (50%), received double weight (2.0), while remaining analytes received single weight (1.0) [Table S3]. Heatmap samples were scored on a scale ranging from 0 to 100, by adding the weights for each discriminatory cytokine above threshold and setting the maximum possible score to 100. A final score-threshold was chosen based on MCC optimization against the heatmap labels, representing the decision boundary between platform responders and nonresponders.

#### Quality control (QC) assessment

Quality control assessment of each sample considered both sample quality and assay quality. Sample quality assessment consisted of two criteria: baseline IL1-ra secretion greater than or equal to 10 pg/ml/hr or Observation Quality passed count (OQPC) greater than or equal to 15. OQPC counts the number of analytes (out of 46) per sample for which time course cumulative concentrations are increasing and the final concentration measured is above the LLOQ. Failure of both criteria was associated with minimal cytokine enrichment and a low elive index, suggesting poor sample quality (Figure S2A). Assay Quality assessment considered cytokines exhibiting significant decreases in secretion rates in the treatment phase relative to control phase (modified-z delta < -2). Samples exhibiting secretion deceleration for 10 or more of the 45 cytokines (excluding EGF and FGF basic, which frequently measure below LLOQ) were flagged as failing assay QC (Figures S2C-D). A sample that failed sample QC or assay QC, but was not a platform responder, was flagged as QNS.

#### Clinical correlation analysis

Concordance between elive index predictions and clinical outcomes was determined using a predefined framework that accounted for treatment type and treatment overlap. For each patient, the highest elive index observed across all corresponding replicates was used for comparison with the clinical response. For treatment matching specimens, any observed clinical outcome was considered a Tier 1 Validation test of the platform prediction (Table S9). PD-1 and PD-L1 inhibitors were considered a treatment match, given their shared mechanism of action along the PD-1/PD-L1 signaling axis. In partial treatment matching cases where ICI was administered in combination with chemotherapy or other agents, clinical progression was considered a Tier 1 Validation test of the elive index prediction as the clinical progression to the matched ICI component was definitive. On the other hand, in partial treatment matching cases, clinical response could be attributed to the unmatched component. For platform responders, the ex vivo prediction to the matched ICI component was considered testable by the observed clinical response (Tier 2 Validation, Figure 1). For platform nonresponders, the clinical response was assumed to be completely attributable to the unmatched component and did not constitute a test of the elive index prediction.

A weighted binary logistic regression model was constructed to determine statistical significance in the association between the elive index and clinical response. The model was used to predict a two-category clinical outcome: disease control (stable disease, pathological complete response, partial response, or complete response) vs progressive disease. True validation samples were assigned a weight of 1.0, Tier 2 Validation samples were assigned a weight of 0.5, reflecting the approximate hazard ratio for overall survival with pembrolizumab plus chemotherapy versus chemotherapy alone reported in KEYNOTE-189 and pooled NSCLC analyses^30^. Additionally, sensitivity analyses were performed by varying partial-match weights across a broad range (0.0–1.0). Statistical analysis was performed using JMP (v19) software.

#### PFS and TTP analysis

PFS was defined as the time from ICI initiation to the first documented radiographic progression or date of death, whichever is earlier. TTP was defined as the time from ICI initiation to the first documented radiographic progression.

Patients alive on ICI therapy and progression-free at the data cutoff of April 13, 2026 were censored at the date of last evaluable progression-free assessment, defined as the latest last evaluable imaging scan showing non-progression, ICI end-of-treatment date or last clinical follow-up. Patients without at least one evaluable post-baseline disease assessment were excluded. PFS Kaplan-Meier analysis was restricted to patients receiving ICI monotherapy or ICI-ICI combination therapy. Comparison between elive responders and non-responders was performed using the log-rank test. Individual patient trajectories across the full IO cohort (including IO-chemotherapy and IO-other regimens) were included in TTP analysis. All clinical analyses were performed in JMP (v19) software.

## Key Resources Table

*This table lists key reagents, biological samples, deposited data, software, and resources used in this manuscript. Where Research Resource Identifiers (RRIDs) are available, they are provided. Resources marked “Proprietary; available under MTA” may be requested from Elephas Biosciences via the Lead Contact*.

| REAGENT or RESOURCE | SOURCE | IDENTIFIER |
| --- | --- | --- |
| **Antibodies** |  |  |
| Pembrolizumab biosimilar (anti–PD-1, IgG4) | BioXcell | Cat# SIM0010; RRID: AB_3084793 |
| Ipilimumab biosimilar (anti–CTLA-4, IgG1) | BioXcell | Cat# SIM0004; RRID: AB_3084787 |
| Atezolizumab biosimilar (anti–PD-L1, IgG1) | BioXcell | Cat# SIM0009; RRID: AB_3084792 |
| Human IgG4 isotype control | BioXcell | Cat# CP147 |
| Human IgG1 isotype control (for atezolizumab arm) | BioXcell | Cat# CP171 |
| Human IgG1 isotype control (for ipilimumab arm) | BioXcell | Cat# CP174 |
| Mouse anti–smooth muscle actin | Cell Marque | Cat# 202M |
| Rabbit anti-CD3 | Ventana Medical Systems | Cat# 790-4341 |
| Mouse anti-CD68 | Ventana Medical Systems | Cat# 790-2931 |
| Mouse anti–pan-cytokeratin (PanCK) | Ventana Medical Systems | Cat# 760-2595 |
| Goat anti-rabbit IgG HRP polymer | Vector Laboratories | Cat# MP-7451 |
| OmniMap anti-mouse HRP | Ventana Medical Systems | Cat# 760-4310 |
| **Biological samples** |  |  |
| Human tumor biopsies (core needle or forceps), 228 patients across 10+ solid tumor types | Multiple US clinical sites (see Author Affiliations); biobank partners | IRB approval per protocol; ClinicalTrials.gov NCT05478538, NCT05520099, NCT06349642, NCT07327489 |
| **Chemicals, peptides, and recombinant proteins** |  |  |
| elive Gel hydrogel (UV-polymerizable, for LTF encapsulation) | Elephas Biosciences | Proprietary; available under MTA |
| Phosphate-buffered formalin, 10% | Standard histology supplier | N/A |
| Culture medium (LTF cell culture) | See Methods | N/A |
| **Critical commercial assays** |  |  |
| Human XL Cytokine Luminex Performance Assay Premixed Kit (30-plex) | R&D Systems / Bio-Techne | Cat# FCSTM18B-30 |
| Custom Luminex Performance Assay (17-plex, configured for elive platform) | R&D Systems / Bio-Techne | Cat# CUSTOM-LXSA-H-17 |
| **Deposited data** |  |  |
| De-identified patient cytokine profiling data and clinical outcomes | This paper | [to be uploaded as supplemental file with acceptance] |
| elive index source code (feature thresholds, weights, scoring) | This paper | Available upon reasonable request |
| **Software and algorithms** |  |  |
| JMP Statistical Software, version 19 | SAS Institute Inc., Cary, NC | https://www.jmp.com; RRID: SCR_014242 |
| Python, version 3.x | Python Software Foundation | https://www.python.org; RRID: SCR_008394 |
| Optuna hyperparameter optimization framework, version 3.6.1 | Akiba et al. 2019 | https://optuna.org; RRID: SCR_022939 |
| scikit-learn (machine learning library) | Pedregosa et al. 2011 | https://scikit-learn.org; RRID: SCR_002577 |
| SciPy (cumulative distribution transform implementation) | Virtanen et al. 2020 | https://www.scipy.org; RRID: SCR_008058 |
| Modified Voabil Classifier (MVC) — adapted scoring framework | This paper; adapted from Voabil et al. 2021 | See Methods; code available upon request |
| elive index — binary cytokine-based response classifier | This paper | See Methods and Tables S2 and S3 |
| **Other** |  |  |
| NanoCool biothermal shipping container (maintains −2 °C to 10 °C) | Peli BioThermal | Cat# 2-85225 |
| Automated proprietary biopsy cutting device (300-µm LTF sections) | Elephas Biosciences | Proprietary; described in Ramasubramanian et al. 2026 |
| 24-well tissue culture plates | Standard supplier | N/A |

## Lead Contact

Hinco J. Gierman, PhD, Chief Scientific Officer, Elephas Biosciences, 1 Erdman Place, Madison, WI 53717.

## Materials Availability

This study did not generate new unique reagents. The elive platform, elive™ Gel, and associated proprietary reagents are available from Elephas Biosciences u. Requests should be directed to the Lead Contact. Commercial antibodies and assay kits are listed above with vendor and catalog numbers. Patient tumor specimens are not available for distribution due to limited tissue quantities and the terms of patient informed consent.

## Data and Code Availability

De-identified patient cytokine profiling data and clinical outcomes will be attached in supplement with publication. Original code for the elive index, including feature thresholds, weighting, and scoring functions, will be made available upon request. Any additional information required to reanalyze the data reported in this paper is available from the Lead Contact upon request.

## Supplemental Figures

**Supplemental figure 1.**
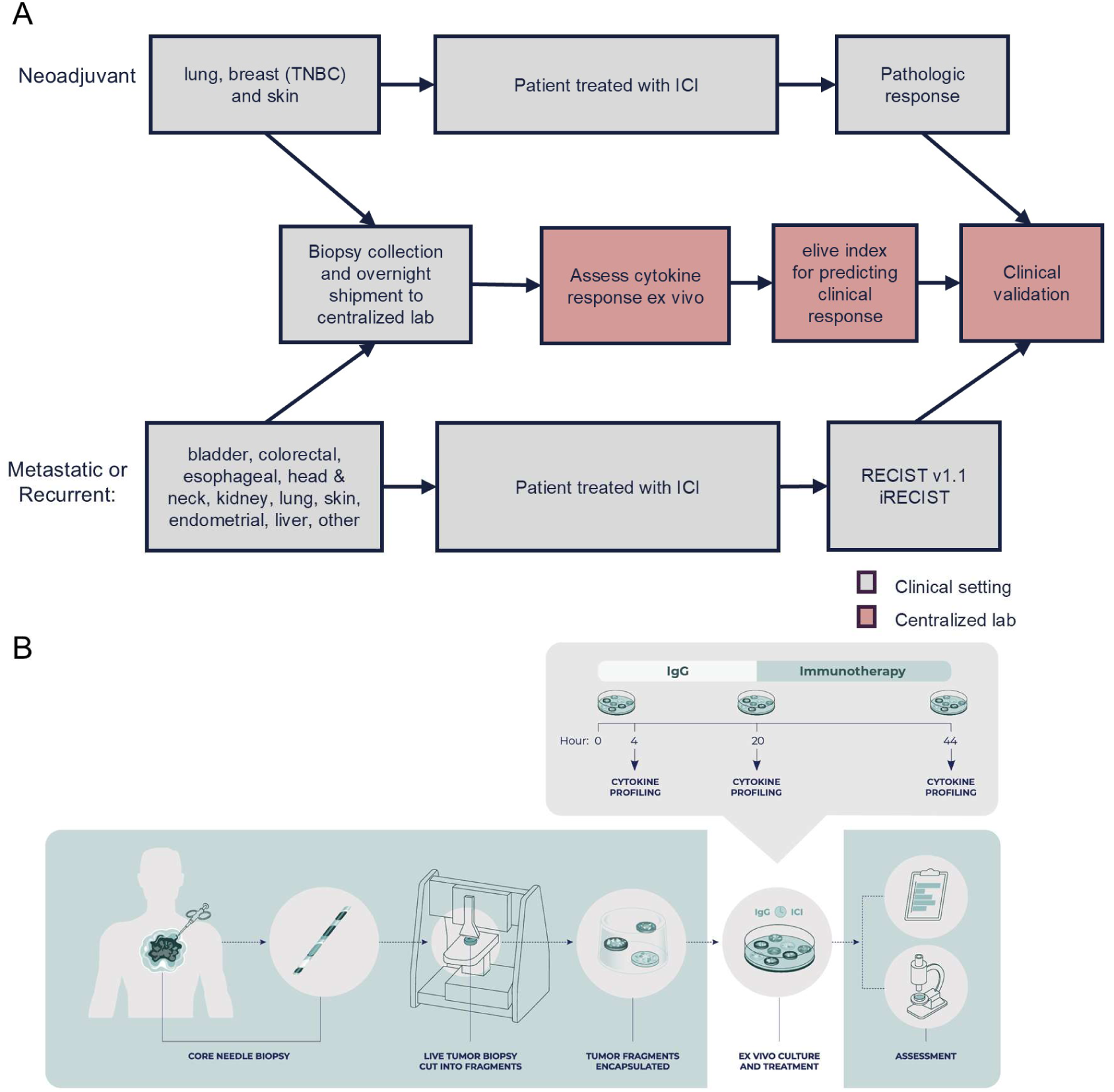
Overview of ongoing observational clinical trials assessing prediction of clinical outcomes with elive. **(A)** Specimens are collected from patients with solid tumors in the neoadjuvant and advanced disease settings, where immunotherapy is commonly administered, and are processed using the platform. Biopsy specimens are evaluated ex vivo for cytokine response to ICI treatment to generate the elive index. Patients subsequently receive standard-of-care therapy as determined by the treating physician, and clinical response is assessed by pathological response in the neoadjuvant setting and by RECIST v1.1 /iRECIST/ real world evidence criteria in metastatic or recurrent disease. The primary objective of these observational trials is to validate the ability of the elive index to predict patient clinical response to ICI therapy. **(B)** The platform integrates an automated tissue-cutting instrument, a proprietary hydrogel, and a sequential treatment methodology to enable functional assessment of ICI response in patient biopsy specimens.

**Supplemental Figure 2.**
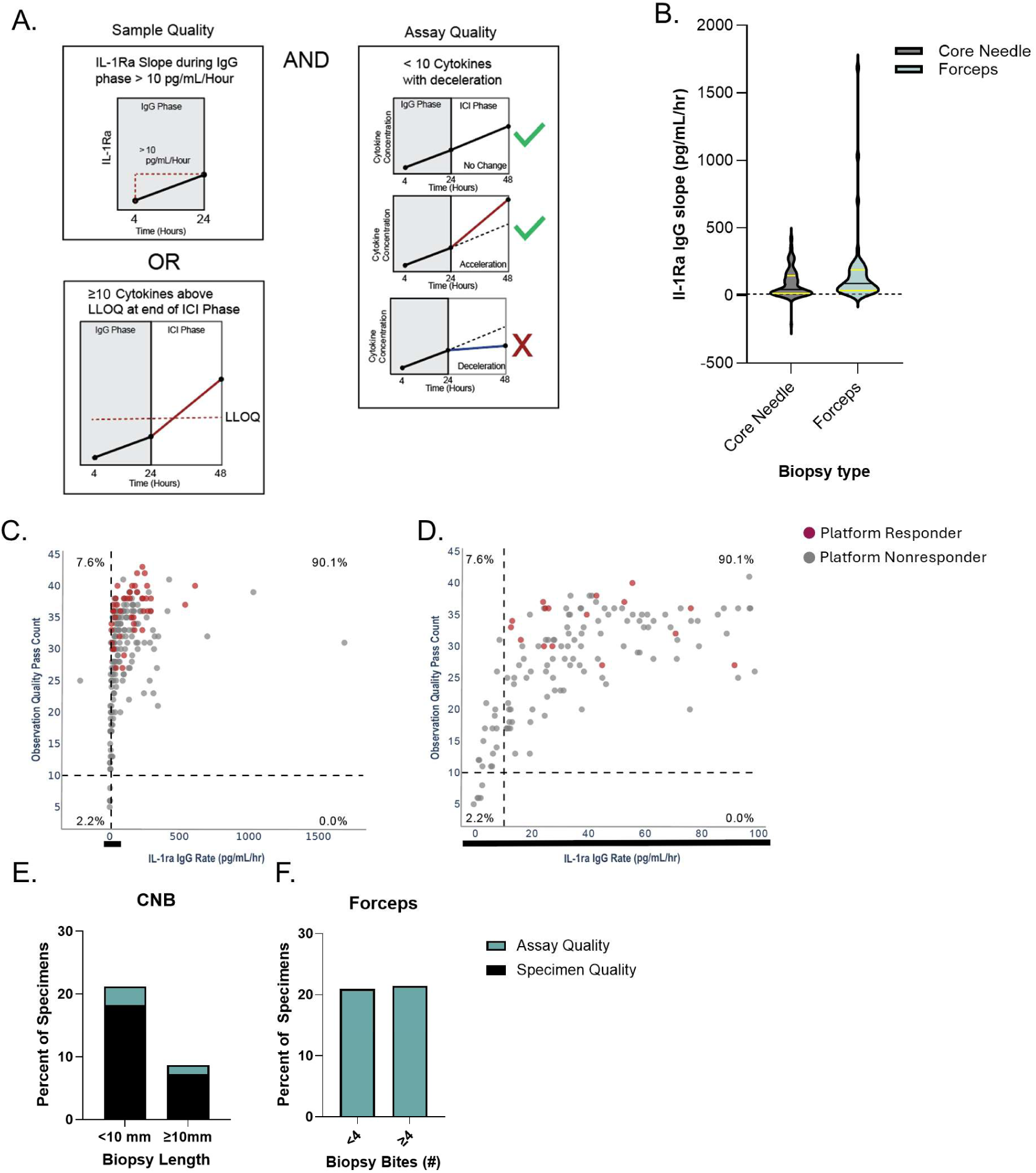
Quality control metrics remove samples based on specimen and assay quality. **(A)** Sample quality criteria (left) requires that the IL-1Ra secretion during the lgG control phase must exceed 10 pg/ml/hour and that >15 out of 46 cytokines must reach above the LLOQ of the cytokine assay at the end of the ICI phase to be considered valid. Sample replicates failing either of these criteria are excluded. Assay quality criteria (right) require that the slope of cytokine induction during the treatment phase must be either the same or increased compared to the lgG phase. Sample replicates exhibiting stable secretion or expected acceleration over time pass QC, whereas replicates showing deceleration are excluded. **(B).** Violin plots show IL-1Ra lgG induction rates stratified by biopsy type (CNB vs. forceps). **(C)** Relationship between IL-1Ra lgG secretion rate and the percentage of cytokines above the LLOQ at the end of the ICI phase, with the QC thresholds indicated by the dashed lines. **(D)** Zoomed-in view of panel C highlighting specimens within the lower range of IL-1Ra lgG secretion rates. Percentages indicate the fraction of specimens in each quadrant. **(E-F)** Distribution of QC failure types are presented for CNBs by biopsy length **(E),** and forceps biopsies by number of biopsies **(F).**

**Supplemental Figure 3.**
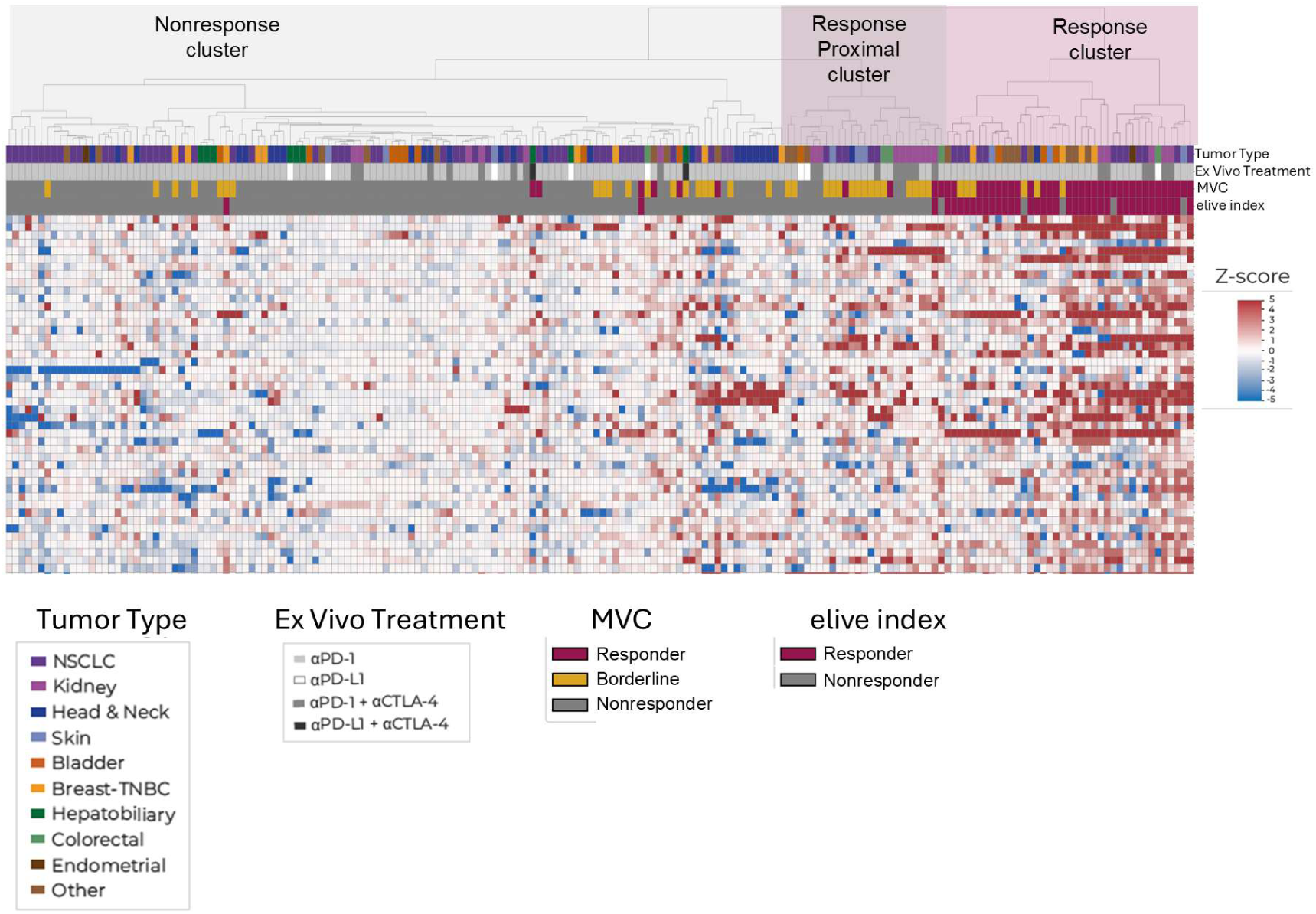
Unsupervised hierarchical clustering of ex vivo cytokine responses to ICI treatment. Heatmap shows z-scores of cytokine measurements across patient samples (n=186 samples from 124 patient tumors) following ex vivo ICI treatment, with hierarchical clustering identifying a response cluster comprising 25% of patients (n= 38 samples from 31 patient tumors) and a nonresponse cluster including a proximal response cluster. Heatmap color reflects modified z-scores of the delta in cytokine production rates between the ICI and lgG treatment phases. Annotation bars indicate cancer type, ex vivo treatment, MVC and elive index.

**Supplemental Figure 4.**
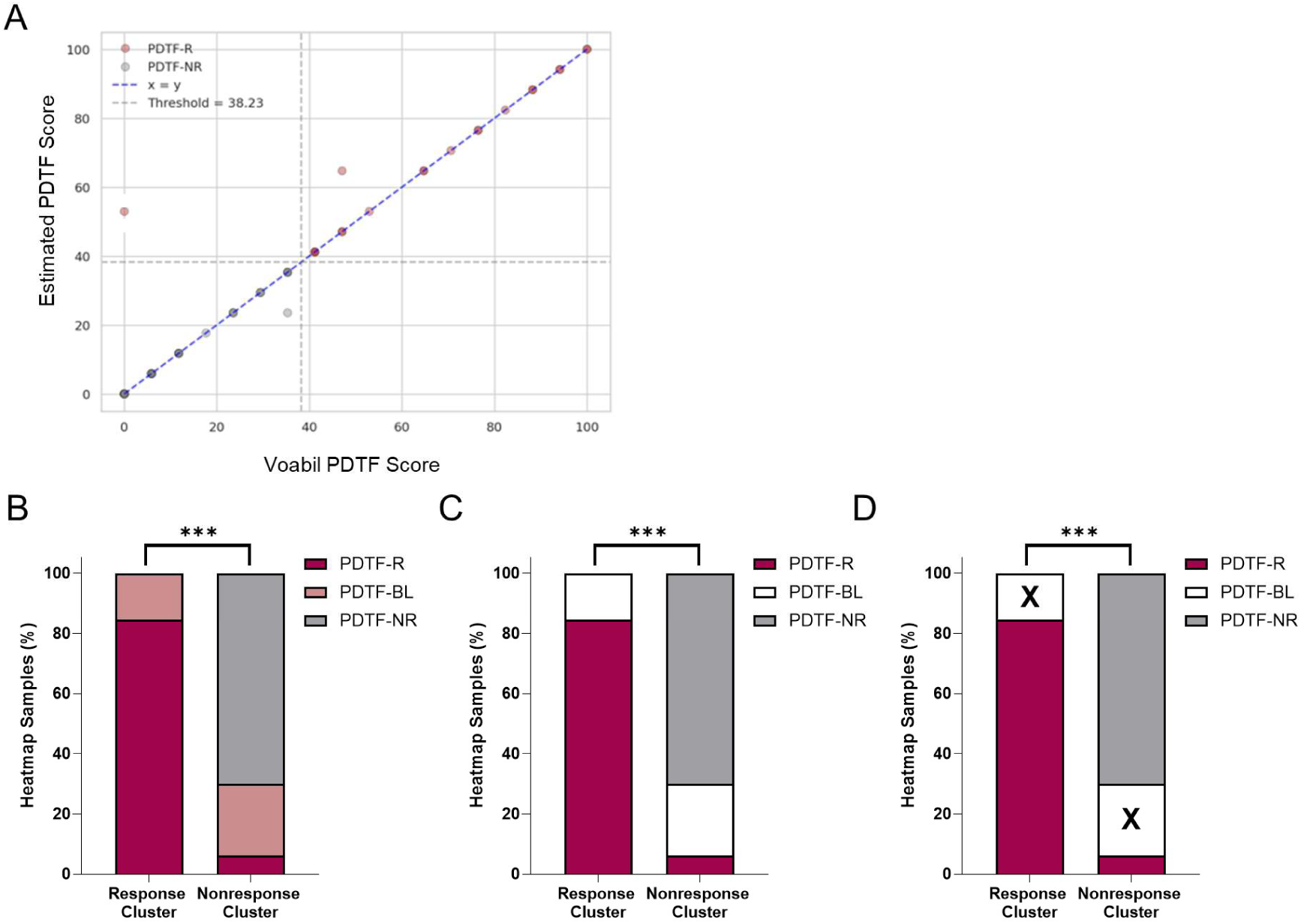
Adaptation of the Voabil et al. PDTF scoring algorithm to LTF platform data validates the use of hierarchical clustering for prediction of immune response on CNBs. **(A)** Comparison of estimated PDTF scores with published Voabil et al. PDTF scores, demonstrating accurate reproduction of the original scoring algorithm following re-engineering and adaptation. The dashed line indicate the responder threshold defined in the Voabil et al. PDTF framework. **(B-D)** Distribution of PDTF-based response classifications within heatmap-defined response and non-response clusters. Most samples within the response cluster are classified as predicted responders (PDTF-R) to aPD-1 using the re-engineered PDTF scoring algorithm, while samples in the non-response cluster are predominantly nonresponders (PDTF-NR). Regardless of the placement of the borderline specimens (PDTF-BL) in analyses with PDTF-R (B), PDTF-NR (C) or exclusion (D) enrichment of the PTDF-Rs in the response cluster was statistically significant.*** P<1 x 10^-16^

**Supplemental Figure 5.**
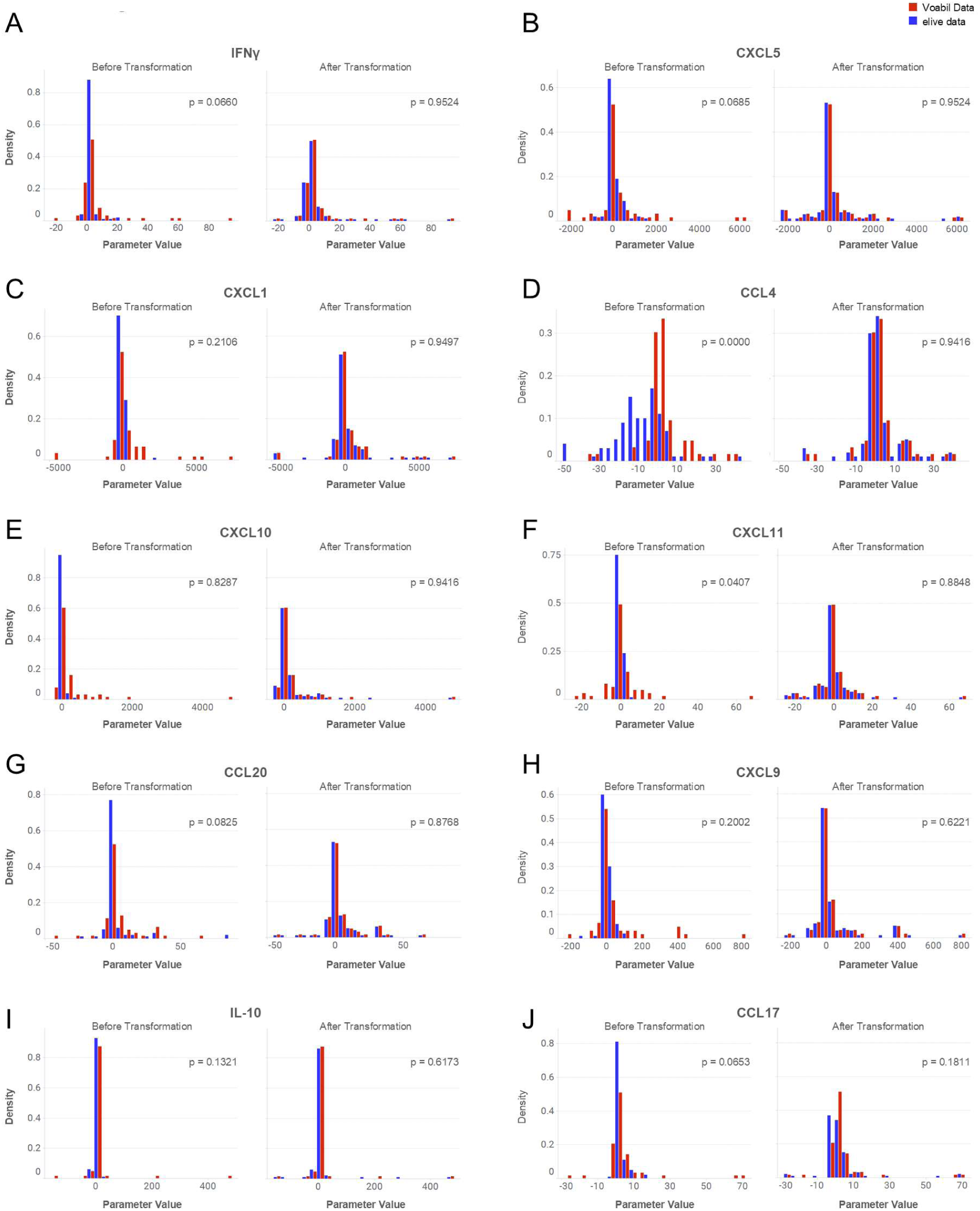
Quantile remapping enables alignment of LTF platform cytokine data with the Voabil et al. PDTF scoring framework. (A-J) Density distributions for individual cytokines included in the re-engineered Voabil et al. PDTF scoring algorithm are shown for LTF platform data (blue) and Voabil et al. reference data (red), before and after quantile remapping. Quantile remapping reshapes and rescales LTF-derived cytokine measurements into the Voabil et al. PDTF data space, enabling direct application of the published Voabil et al. PDTF scoring algorithm to elive platform outputs. Across the cytokines evaluated, 9 of 10 cytokine distribution mappings demonstrate excellent concordance between the LTF platform and Voabil et al. datasets following transformation.

**Supplemental Figure 6.**
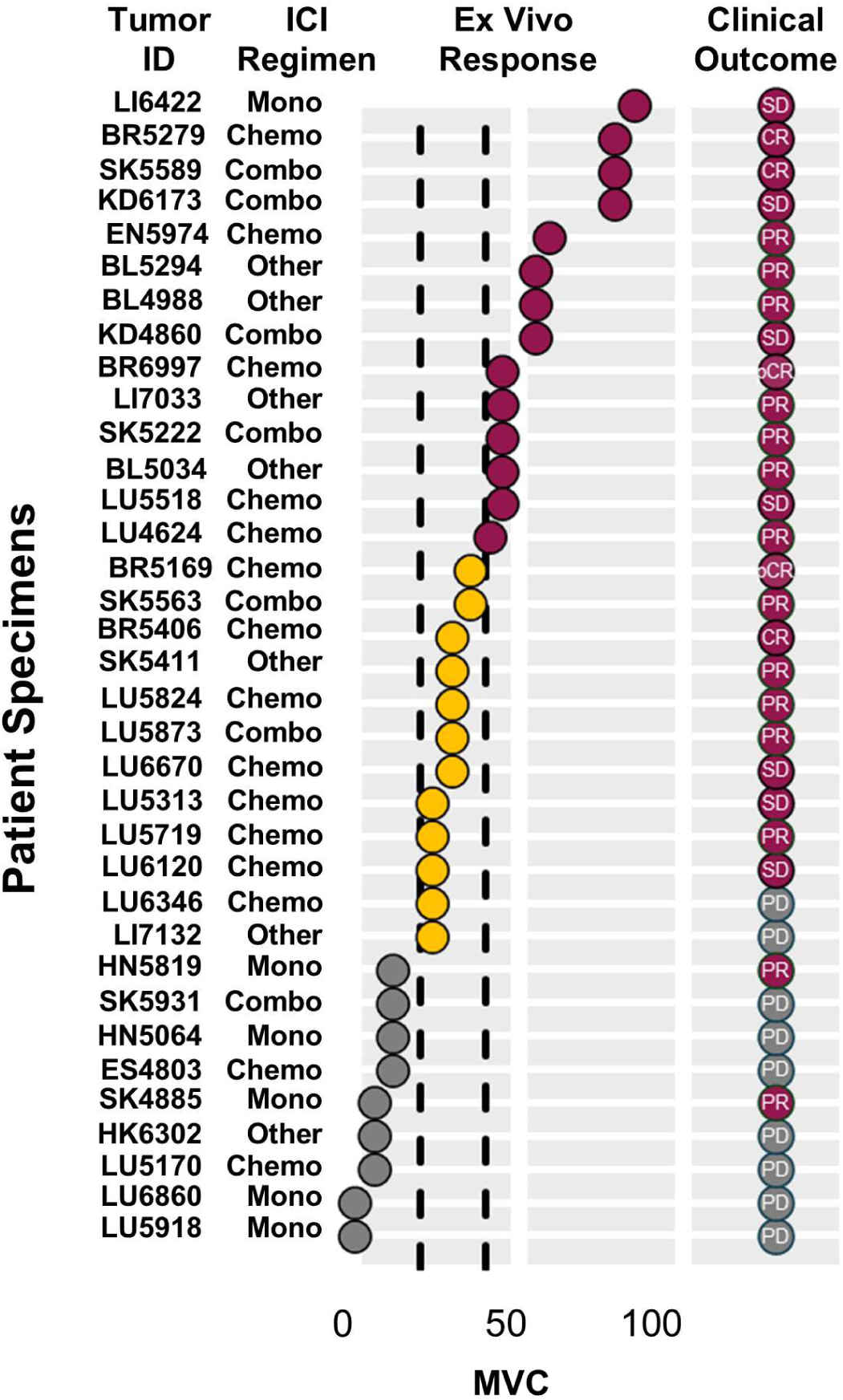
Modified Voabil Classifier (MVC) accurately predicts clinical response to immune checkpoint inhibitors in 21/23 patients where a determinate responder/nonresponder call was possible. MVC Response Score and accompanying clinical response (best overall response) for all specimens and patients where there was an ICI treatment match between the ex vivo and clinical settings. Treatment match was classified into 3 categories (1) perfect match= ICI mono or combination therapy target(s) were matched between the ex vivo and clinical settings, (2) partial treatment match(*) = ICI monotherapy target ex vivo matches one arm of clinical combination treatment regimen, (3) not applicable = MVC Response Score falls within borderline region. Specimen IDs and clinical treatment regimens are reported on the y-axis. Broken black lines represent MVC Score thresholds used to define MVC predicted responder (2:38.23; red dots), MVC predicted nonresponder (:520.58; gray dots) and borderline specimens (>20.58 and <38.32; yellow dots).

**Supplemental Figure 7.**
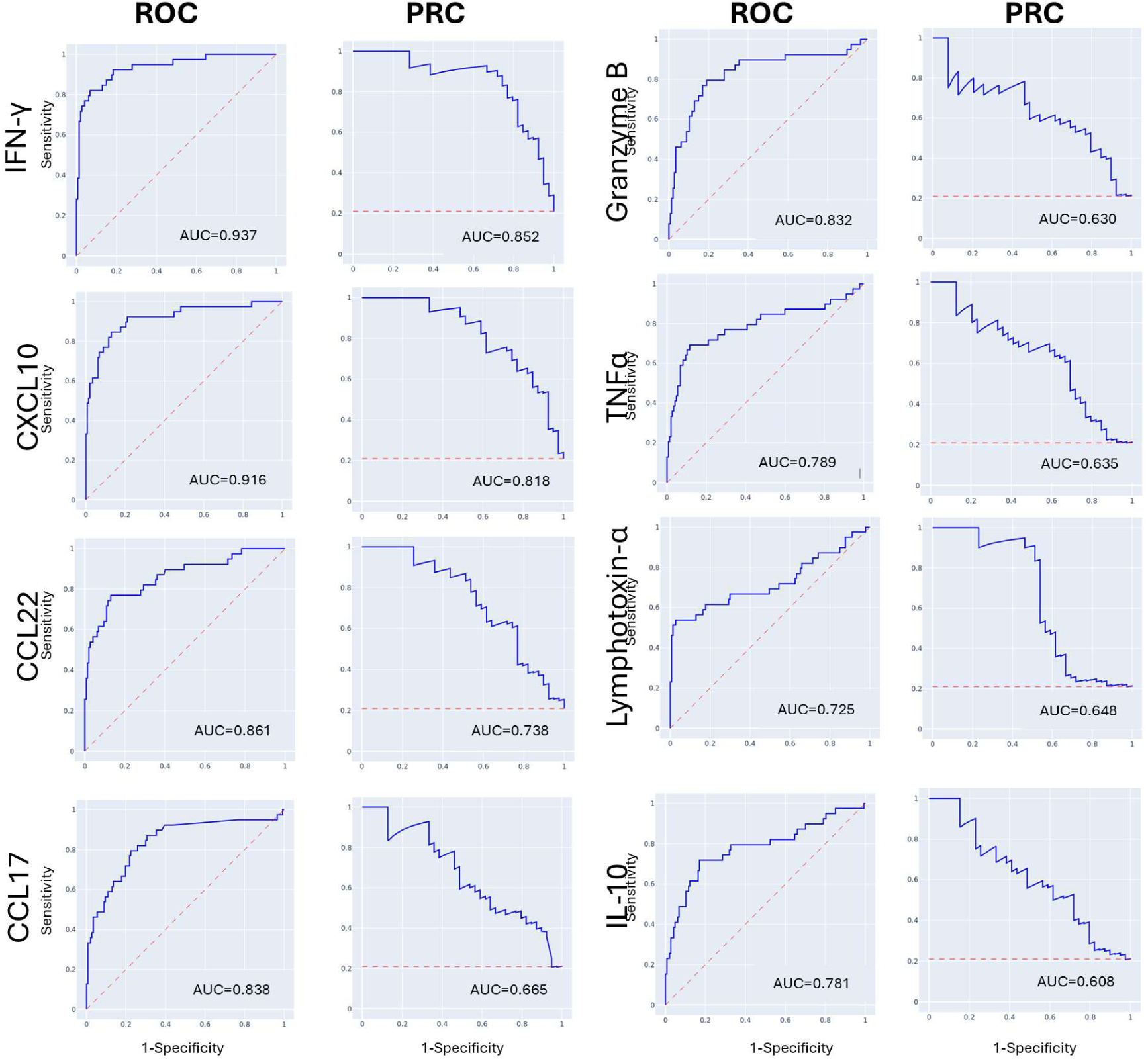
AUROC and AUPRC for the 8 cytokines included in elive index. AUC values are reported for individual ROCs and PRCs across cytokines included in the elive index.

**Supplemental Figure 8.**
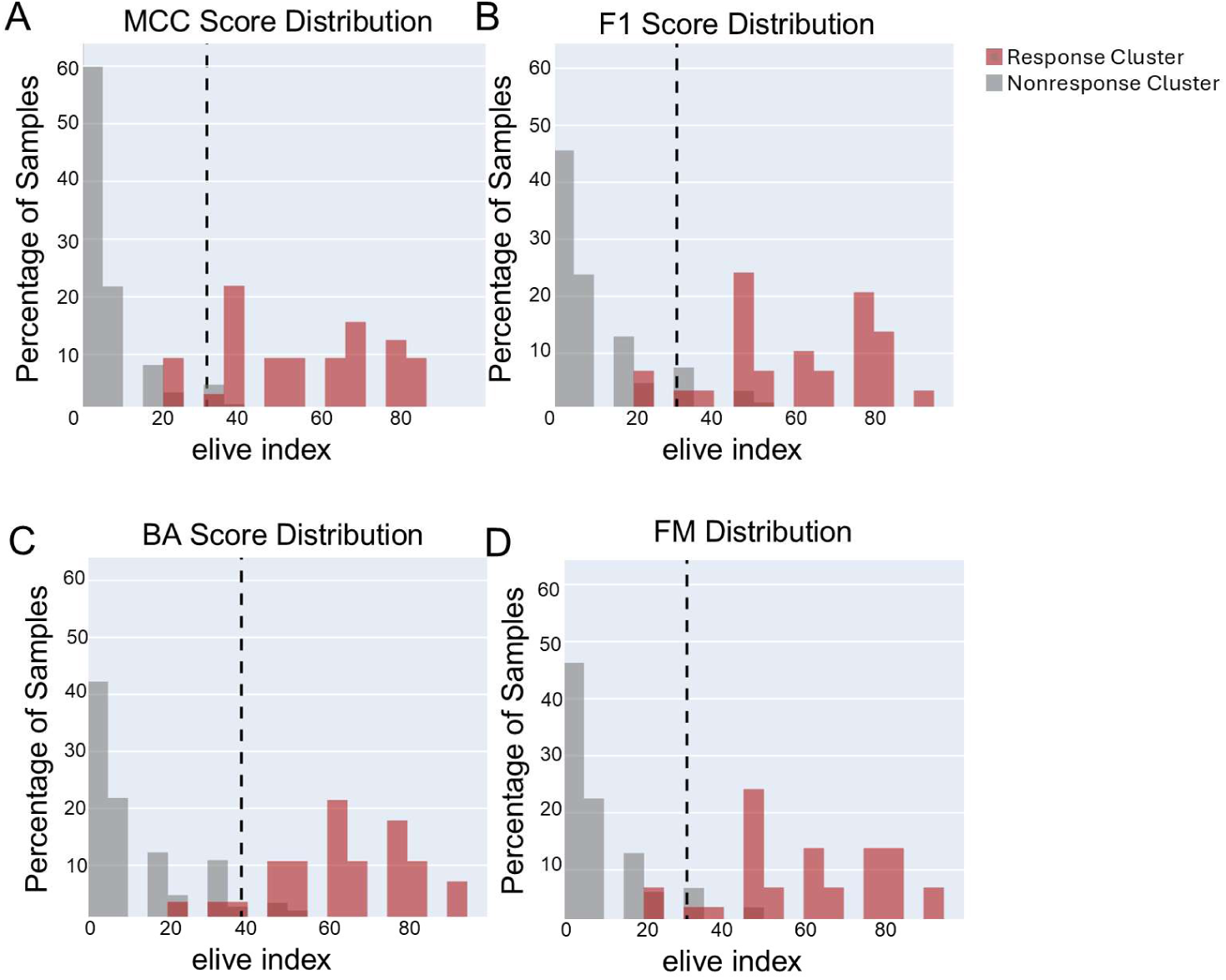
Comparison of thresholding performance metrics used for cytokine-based elive index. Distributions of elive indexes generated using cytokine thresholds optimized by **(A)** Matthews correlation coefficient (MCC), **(B)** F1 score, **(C)** balanced accuracy (BA), and **(D)** Fowlkes-Mallows (FM) distribution. Histograms display the percentage of samples across elive index ranges. The dashed vertical line denotes the score threshold of 100% sensitivity.

**Supplemental Figure 9.**
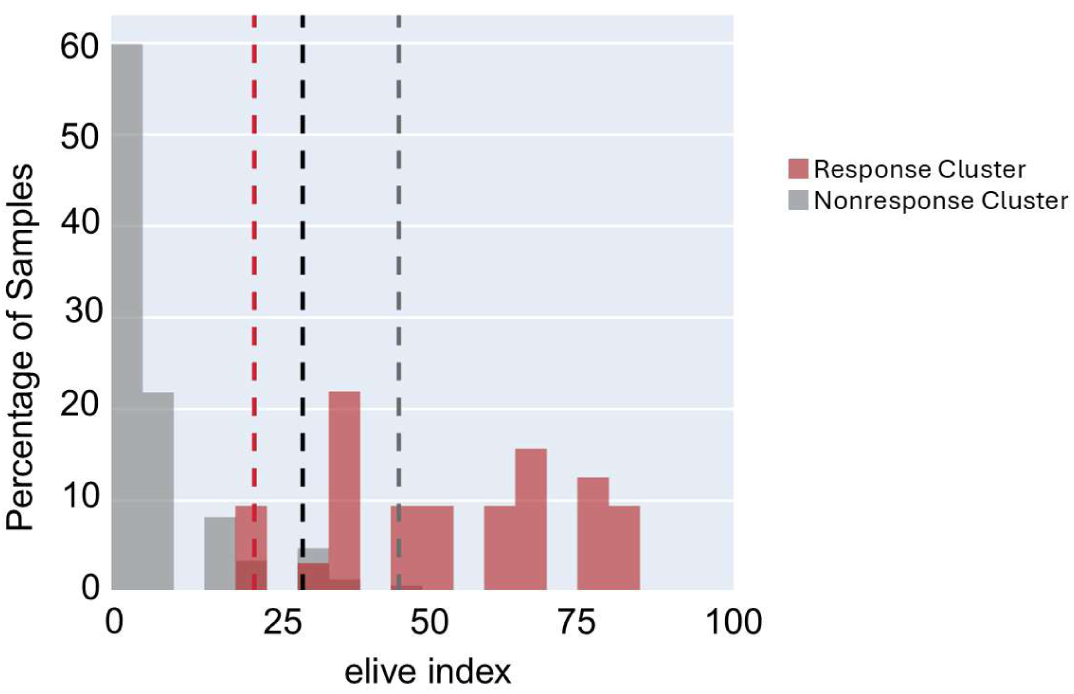
MCC analysis results in the optimal balance between precision and sensitivity for identifying heatmap response samples. Histogram showing the distribution of samples across elive index values, with response-cluster samples shown in red and non-response-cluster samples in gray. Dashed lines indicate candidate thresholds for classifying response versus non-response: black denotes a sensitivity-based threshold, red denotes the MCC-based threshold, and blue denotes the specificity-based threshold.

**Supplemental Figure 10.**
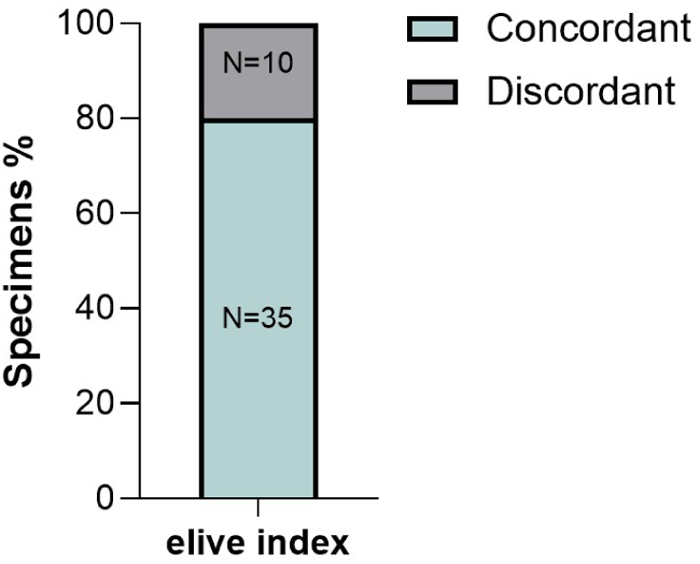
Concordance of elive index across replicate sam ples. Among all specimens in the training dataset with replicate samples assessed, 35 of 45 specimens (78%) show complete concordance of elive index outcomes across all replicates.

**Supplemental Figure S11.**
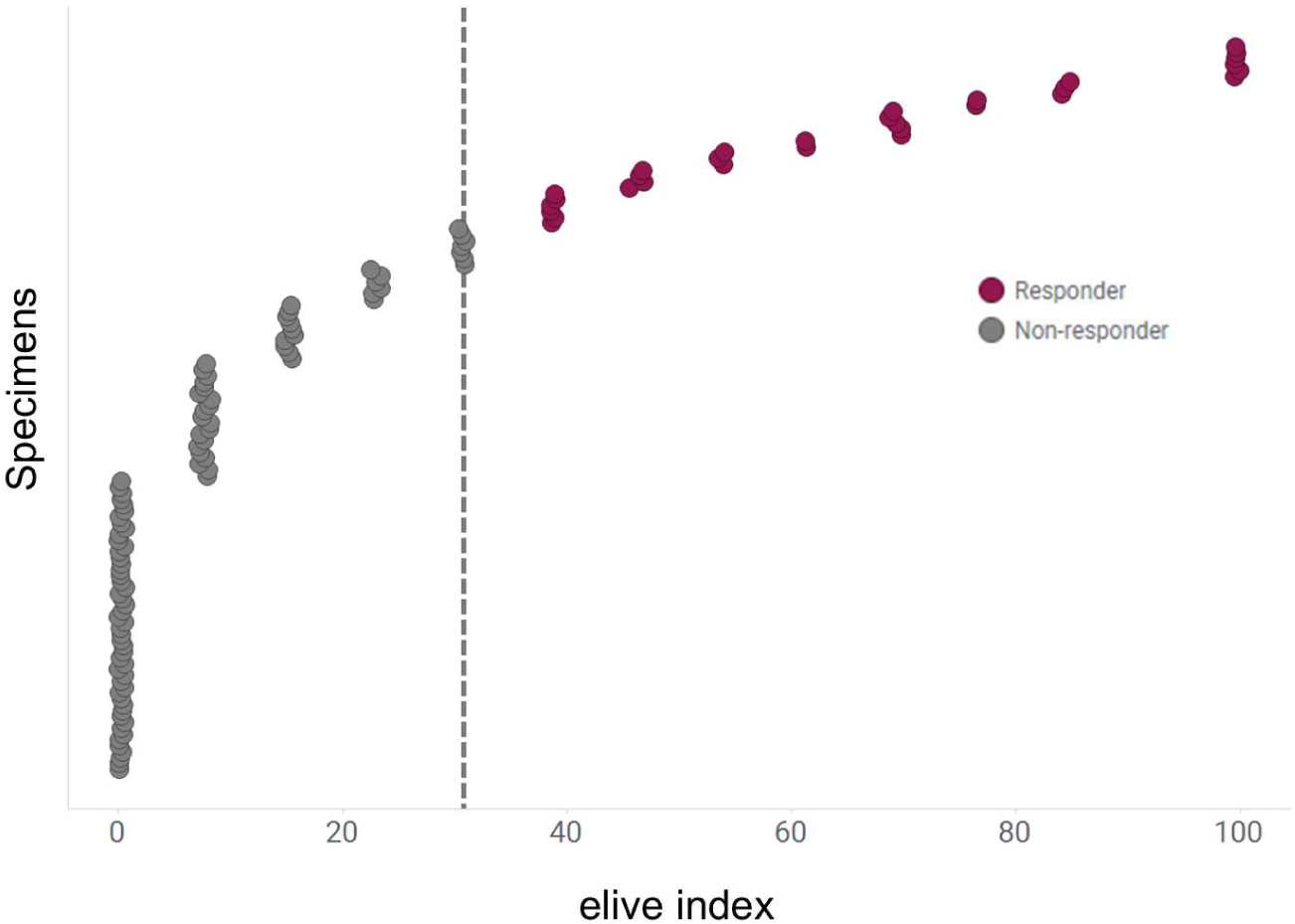
elive index predicts 26% of training specimens are cytokine responders. elive index for all specimens included in training heatmap (n=124). Predicted responders (elive index >30.8) are depicted in red (n=32) and nonresponders (elive index: 530.8) in gray (n=92). For specimens with multiple replicates, the highest elive index is reported. Note application of x-axis jitter to enable visualization of overlapping data points.

**Supplemental Figure 12.**
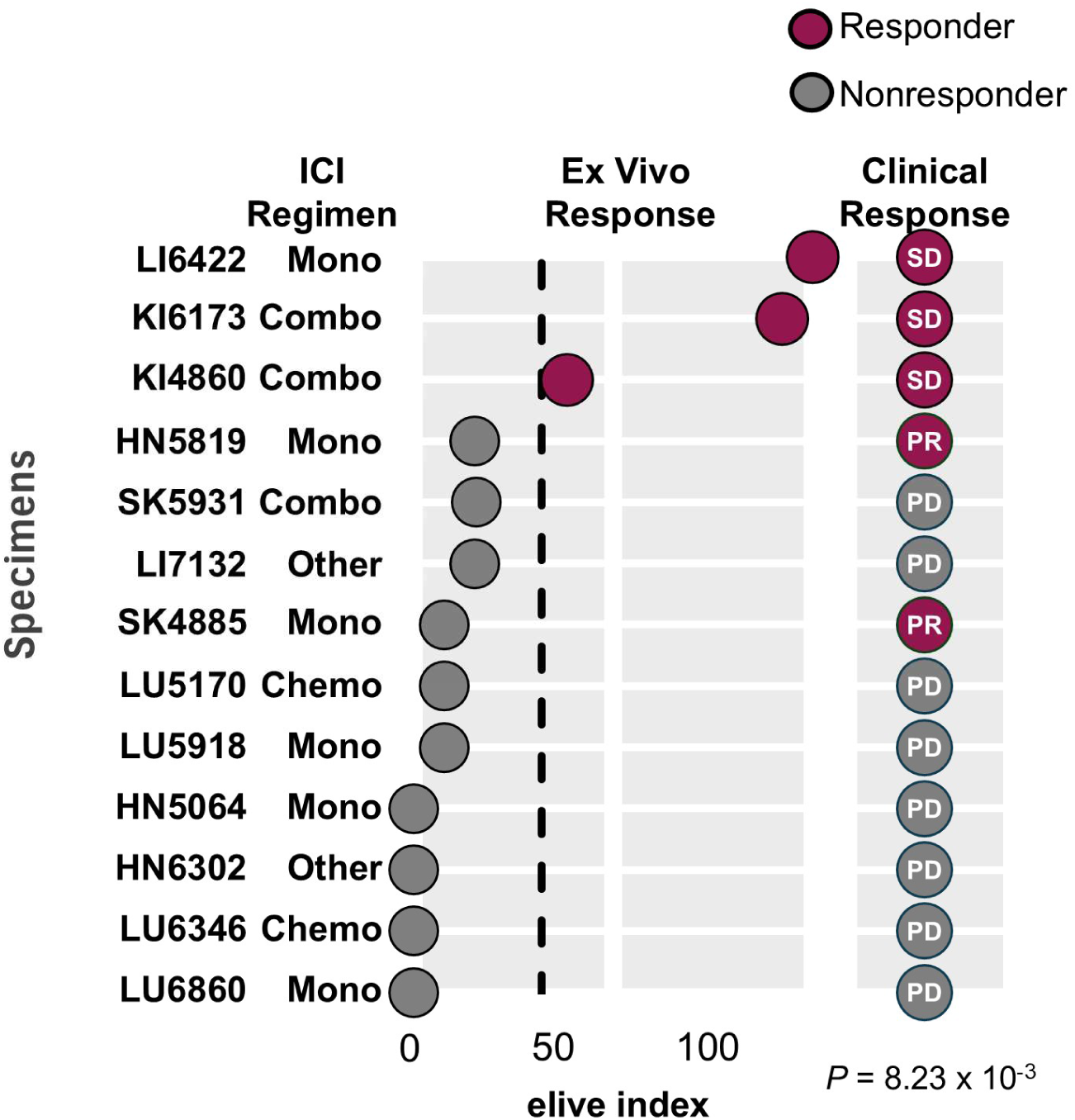
elive index accurately predicts clinical response to ICI in 85% of Tier 1 Validation patients. elive index and accompanying clinical response (best overall response) for all specimens and patients where there was a perfect ICI target match between the ex vivo and clinical settings and those with clinical progression on any ICI-containing combination regimen (ICI + chemo, TKI, ADC or ICI). Specimen IDs and clinical treatment regimens are reported on the y-axis. All specimens were treated ex vivo with aPD-1 except for KI6173, KI4860, SK6874 and SK5931 which were treated with aPD- 1+ aCTLA-4 and LI6422 which was treated with aPD-L1. Broken black line represents elive index threshold used to define platform predicted responders (red dots, elive index >30.8) and nonresponders (gray dots, elive index <30.8).

**Supplemental Figure 13.**
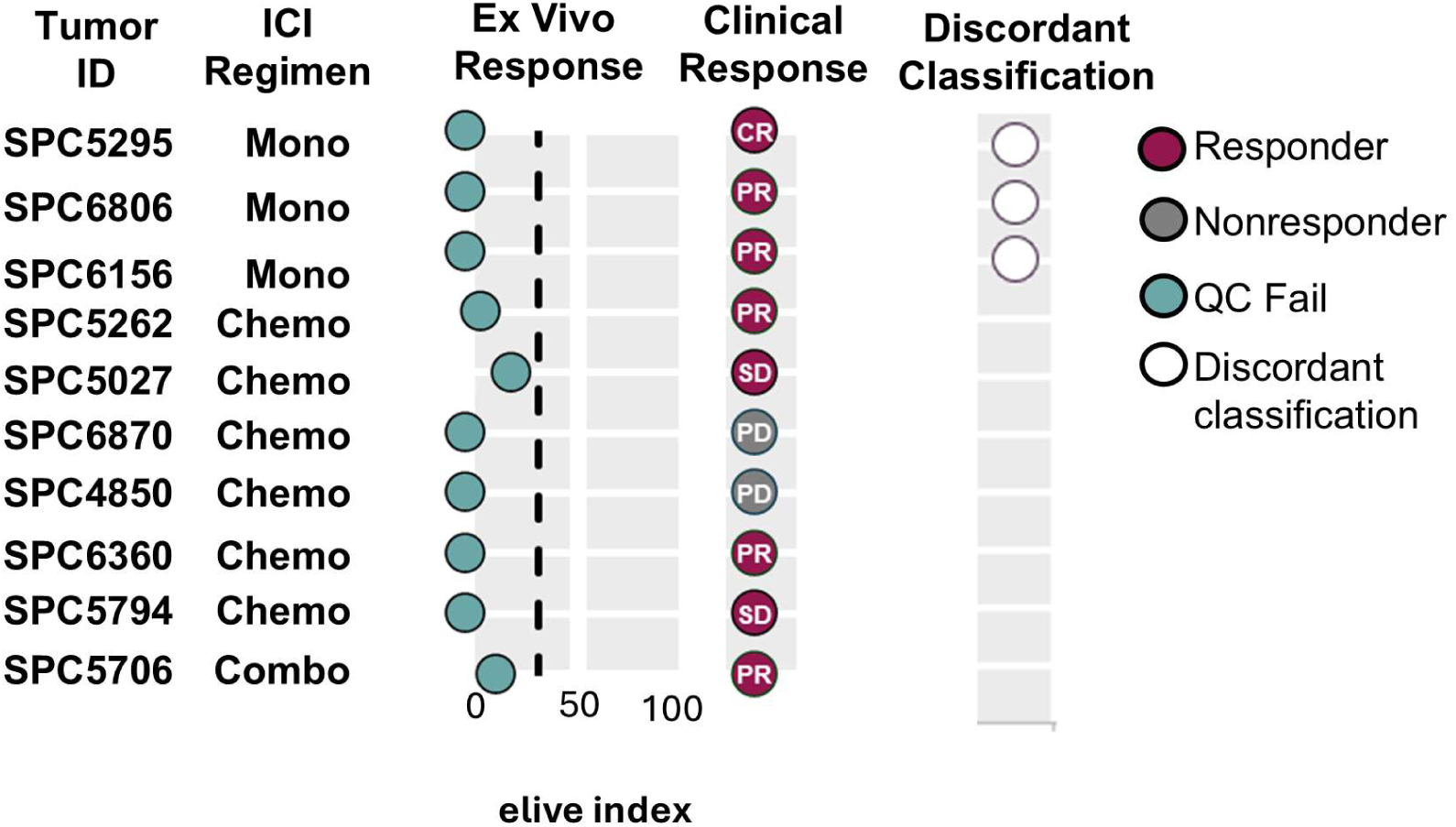
Specimens failing to meet quality control metrics can yield elive indexes that are discordant with clinical outcomes. elive indexes for tumor specimens failing to meet quality control metrics but with an ICI treatment match between the ex vivo and clinical settings and accompanying clinical response data (best overall response). Treatment match was classified into 2 categories (1) perfect match = ICI mono or combination therapy target(s) were matched between the ex vivo and clinical settings, (2) partial treatment match (*) = ICI monotherapy target ex vivo matches one arm of clinical combination treatment regimen. Specimen IDs and clinical treatment regimens are reported on the y-axis. All specimens were treated ex vivo with aPD-1. Broken black line represents elive index threshold used to define platform predicted responders (elive index >30.8) and nonresponders (elive index <30.8) for specimens passing QC. Clinical response is reported as CR = complete response (red), PR = partial response (red) and PD = progressive disease (gray).

**Figure S14.**
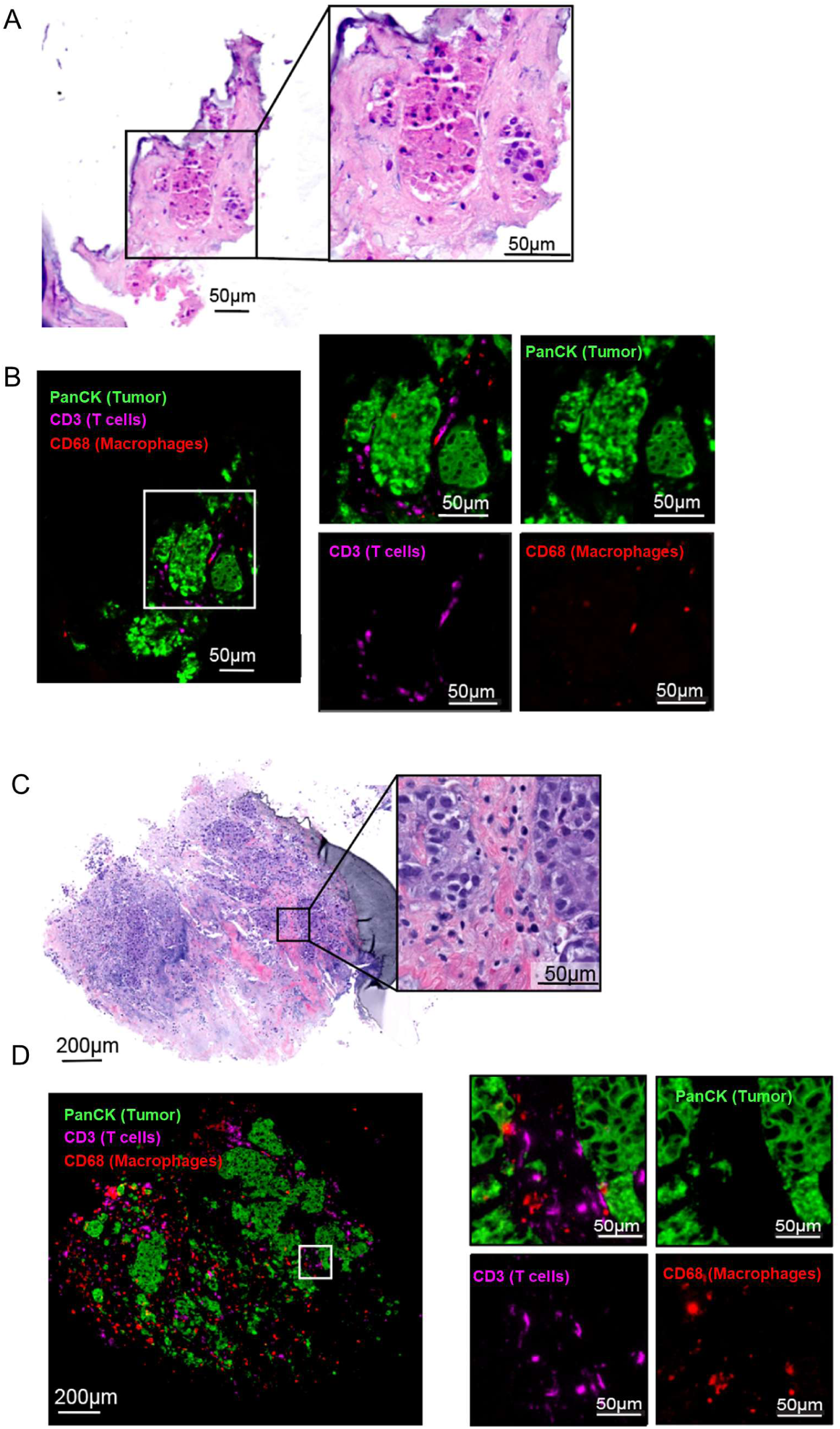
Histology from nonresponding and responding specimens demonstrates preservation of immune cells and tumor architecture. **(A)** H&E labeling of representative LTF of nonresponding specimen HN5064. **(B)** mlF labeling of the same LTF, showing low magnification (left) with white inset box indicating the region shown at higher magnification on the right. Higher-magnification panels display the composite image alongside individual channels for PanCK (tumor; green), CD3 (T cells; magenta), and CD68 (macrophages; red). **(C)** H&E labeling of representative LTF of responding specimen TNBC5279. **(D)** mlF labeling of same LTF showing low magnification (left) with white box representing higher magnification on the right with composite as well as individual channels labeling PanCK (tumor, green), CD3 (T cells, magenta), CD68 (macrophages, red)

**Supplemental Table 1.** Biopsy type, size, and sampling distribution processed on the elive platform. The table summarizes the range of biopsy types and gauge sizes successfully processed, including CNBs and forceps biopsies.

| Biopsy Type | N | % |
| --- | --- | --- |
| **CNBs** | 117 | 66.9 |
| 9 G | 1 | 0.9 |
| 14 G | 6 | 5.1 |
| 16 G | 8 | 6.8 |
| 18 G | 83 | 70.9 |
| 20 G | 18 | 15.4 |
| 22 G | 1 | 0.9 |
| **Forceps** | 58 | 33.1 |
| Standard<br>1.9 - 2.4 mm | 56 | 96.6 |
| Small<br>< 1.8 mm | 1 | 1.7 |
| Large<br>> 2.4 mm | 1 | 1.7 |
| **Number of Biopsies**<br>**per Patient** | **CNBs (N)** | **Forceps (N)** |
| 1 | 58 | 1 |
| 2 | 35 | 4 |
| 3 | 16 | 39 |
| 4+ | 8 | 14 |

**Supplemental Table 2.**
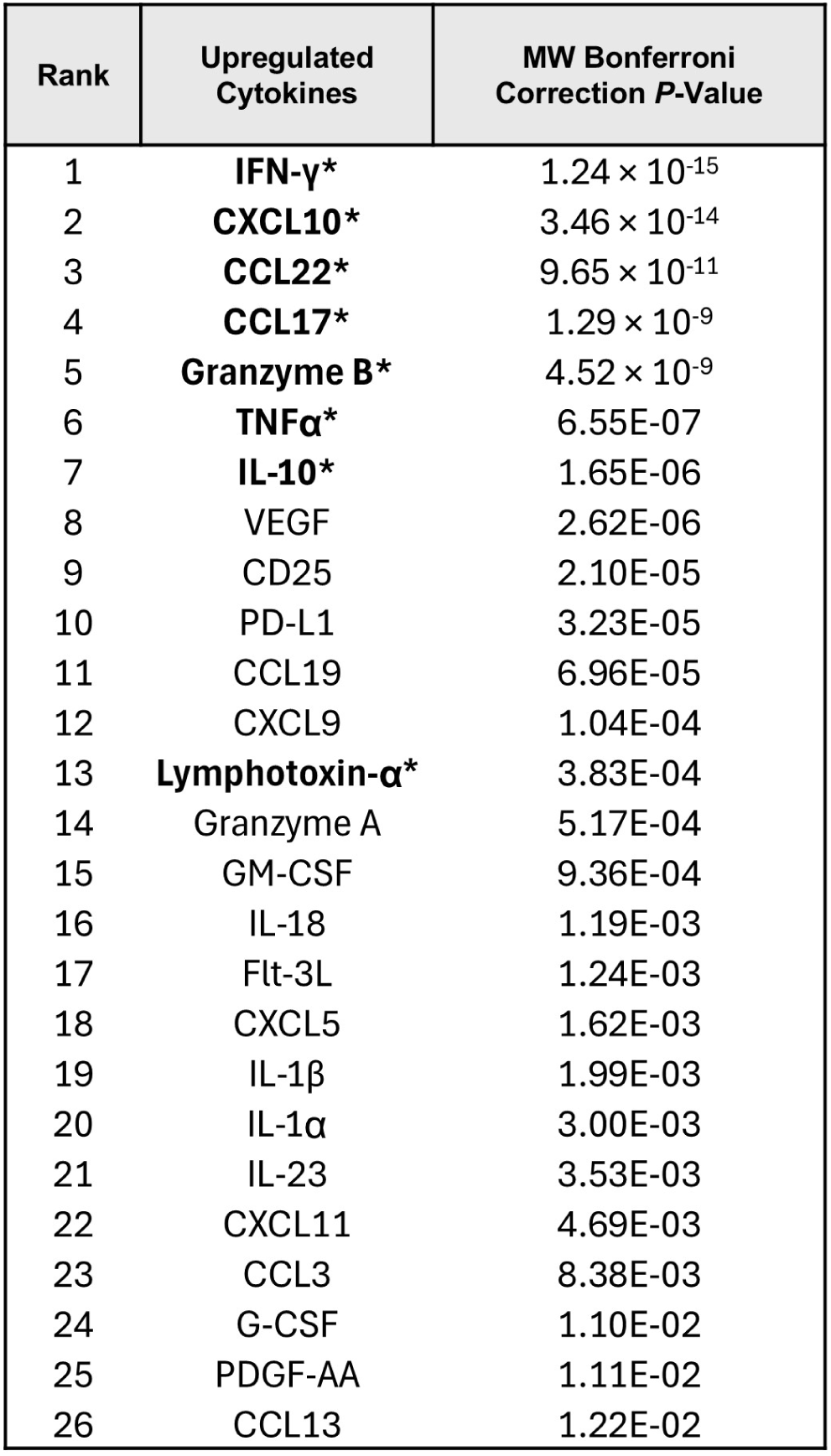
26 cytokines were significantly upregulated in the response cluster. Rank order of 26 significantly upregulated cytokines in heatmap response cluster in comparison to the non-response cluster per Mann-Whitney U test and Bonferroni Correction. Cytokines included in elive index classifier are in bold and annotated with an asterisk.

**Supplemental Table 3.** Cytokines with the greatest discriminative power between heatmap-defined response and non-response clusters. Table lists normalized threshold values and corresponding weights used in the calculation of the elive index, along with sensitivity and specificity metrics describing each analyte’s ability to distinguish between response and non-response clusters.

| Analyte | Normalized threshold | Weight | HM sensitivity | HM specificity |
| --- | --- | --- | --- | --- |
| IFN-γ | 4.7 | 2 | 0.74 | 0.97 |
| CCL22 | 3.2 | 2 | 0.54 | 0.97 |
| CXCL10 | 2.8 | 2 | 0.74 | 0.93 |
| Lymphotoxin-α | 1.5 | 2 | 0.51 | 0.99 |
| TNFα | 0.9 | 2 | 0.59 | 0.93 |
| CCL17 | 8.4 | 1 | 0.46 | 0.97 |
| Granzyme B | 1.2 | 1 | 0.77 | 0.83 |
| IL-10 | 0.4 | 1 | 0.72 | 0.83 |

**Supplemental Table 4.** MCC analysis results in the optimal balance between precision and sensitivity for identifying heatmap response samples. Table summarizing threshold values and associated performance metrics for 3 elive index thresholding models. The optimal model for each metric is highlighted in red. Overall, selecting the elive index threshold based on MCC optimizes the majority of performance metrics compared with thresholds based on sensitivity or specificity alone.

| elive index threshold performance metrics |  |  |  |  |  |
| --- | --- | --- | --- | --- | --- |
| Method | AUC | Sens | Spec | MCC | F1 |
| **MCC** | **0.99** | 0.90 | **0.98** | **0.89** | **0.91** |
| **F1** | 0.98 | 0.92 | 0.95 | 0.83 | 0.87 |
| **BA** | **0.99** | 0.92 | 0.95 | 0.83 | 0.87 |
| **Distance** | 0.98 | **0.95** | 0.90 | 0.77 | 0.81 |

**Supplemental Table 5.** elive index is significantly correlated with clinical response at all weights for Tier 2 Validation specimens. P-value sensitivity analysis using binary logistic regression to determine significance of the relationship between elive index and clinical response using weights varying from 0.0 to 1 for Tier 2 Validation specimens (Tier 1 Validation specimens were assigned a weight of 1). Analysis applies disease control classification of clinical response binning CR, pCR, PR, SD vs. PD for binary classification.

| Tier 2 Validation Weight | *P*-value |
| --- | --- |
| 0 | $8.27 \times 10^{-3}$ |
| 0.1 | $1.97 \times 10^{-3}$ |
| 0.2 | $5.80 \times 10^{-4}$ |
| 0.3 | $1.98 \times 10^{-4}$ |
| 0.4 | $7.66 \times 10^{-5}$ |
| 0.5 | $3.24 \times 10^{-5}$ |
| 0.6 | $1.48 \times 10^{-5}$ |
| 0.7 | $7.18 \times 10^{-6}$ |
| 0.8 | $3.69 \times 10^{-6}$ |
| 0.9 | $1.98 \times 10^{-6}$ |
| 1 | $1.11 \times 10^{-6}$ |

**Table S6.** Table reports relationship between clinical response, elive index outcome and ICI biomarker status for all Tier 1 Validation and Tier 2 Validation specimens where ICI biomarker data were available. Individual characteristics for patients where elive index predicted a different ICI outcome compared to indicated companion diagnostic ICI biomarkers. Blue shading highlights ex vivo and patient treatment matches. Durva = durvalumab; lpi = ipilimumab; Pembro = pembrolizumab; Garb = carboplatin; Pem = pemetrexed; Nivo = nivolumab; 5-FU = 5-Fluorouracil; Ox= oxaliplatin; Cis = cisplatin; Ptx = paclitaxel.

| Patient | Correct Predictor of Response | Clinical Response | elive index Outcome | ICI Biomarker Status | Tumor Type | Clinical Treatments | Ex Vivo Treatment |
| --- | --- | --- | --- | --- | --- | --- | --- |
| HN5819 | ICI Biomarker | Partial Response | Nonresponder | Positive (PD-L1) | Head and Neck | Pembro | αPD-1 |
| ME4885 | ICI Biomarker | Partial Response | Nonresponder | Positive (TMB) | Melanoma | Pembro | αPD-1 |
| LI6422 | elive index | Stable Disease | Responder | Negative (TMB & MSI) | Liver | Durva | αPD-L1 |
| KI6173 | elive index | Stable Disease | Responder | Negative (TMB & MSI) | Kidney (CCRCC) | Ipi + Nivo | αCTLA4 + αPD-1 |
| LU5918 | elive index | Progressive Disease | Nonresponder | Positive (PD-L1) | NSCLC | Pembro | αPD-1 |
| LU6860 | elive index | Progressive Disease | Nonresponder | Positive (TMB) | NSCLC | Pembro | αPD-1 |
| UR5034 | elive index | Partial Response | Responder | Negative (MSI) | Urothelial | Pembro + Enfortumab | αPD-1 |
| LU6120 | elive index | Stable Disease | Responder | Negative (PD-L1, TMB, MSI) | NSCLC | Pembro + Ptx + Carb | αPD-1 |
| HN6496 | elive index | Stable Disease | Responder | Negative (TMB & MSI) | Head and Neck | Nivo + Carb + Ptx | αPD-1 |
| ES4803 | elive index | Progressive Disease | Nonresponder | Positive (MSI) | Esophageal | Nivo + 5-FU + Ox | αPD-1 |
| LU6346 | elive index | Progressive Disease | Nonresponder | Positive (TMB) | NSCLC | Pembro + Ptx + Carb | αPD-1 |
| HN5064 | Both | Progressive Disease | Nonresponder | Negative (PD-L1) | Head and Neck | Pembro | αPD-1 |
| LU5687 | Both | Progressive Disease | Nonresponder | Negative (PD-L1, TMB, MSI) | NSCLC | Durva | αPD-1 |
| ME5931 | Both | Progressive Disease | Nonresponder | Negative (TMB, MSI) | Melanoma | Ipi + Nivo | αCTLA4 + αPD-1 |
| EN7132 | Both | Progressive Disease | Nonresponder | Negative (MMR) | Endometrial | Pembro + levatinib | αPD-1 |
| LU5170 | Both | Progressive Disease | Nonresponder | Negative (PD-L1) | NSCLC | Pembro + Cis + Pem | αPD-1 |
| LU4624 | Both | Partial Response | Responder | Positive (TMB) | NSCLC | Pembro + Carb + Pem | αPD-1 |
| BR5279 | Both | Complete Response | Responder | Positive (PD-L1) | TNBC | Pembro + Carb + Ptx | αPD-1 |
| LU5518 | Both | Stable Disease | Responder | Positive (PD-L1, TMB) | NSCLC | Pembro + Carb | αPD-1 |
| UR5294 | Both | Partial Response | Responder | Positive (TMB) | Urothelial | Pembro + enfortumab | αPD-1 |

**Supplemental Table 7.** Ex vivo treatments. Table summarizing catalog numbers for ICI treatments used ex vivo along with associated controls

| Ex vivo treatment |  | IgG control |  |
| --- | --- | --- | --- |
| Drug | BioXCell Cat # | Drug | BioXCell Cat # |
| αPD-1 | SIM0010 | IgG4 | CP147 |
| αPD-L1 | SIM0009 | IgG1 | CP171 |
| αCTLA-4 | SIM0004 | IgG1 | CP174 |

**Supplemental Table 8.** Multiplex immunofluorescence antibody manufacture details. Table summarizing catalog numbers for antibodies used in immunofluorescence labeling

| Antibody | Manufacturer and Catalog Number |
| --- | --- |
| Actin, smooth muscle | Cell Marque #202M |
| anti-CD3 | Ventana Medical Systems #790-4341 |
| anti-CD68 | Ventana Medical Systems #790-2931 |
| anti-Keratin, pan | Ventana Medical Systems #760-2595 |
| Goat anti-rabbit IgG HRP | Vector Laboratories #MP-7451 |
| Omni-map anti-mouse HRP | Ventana Medical Systems #760-4310 |

**Supplemental Table 9.**
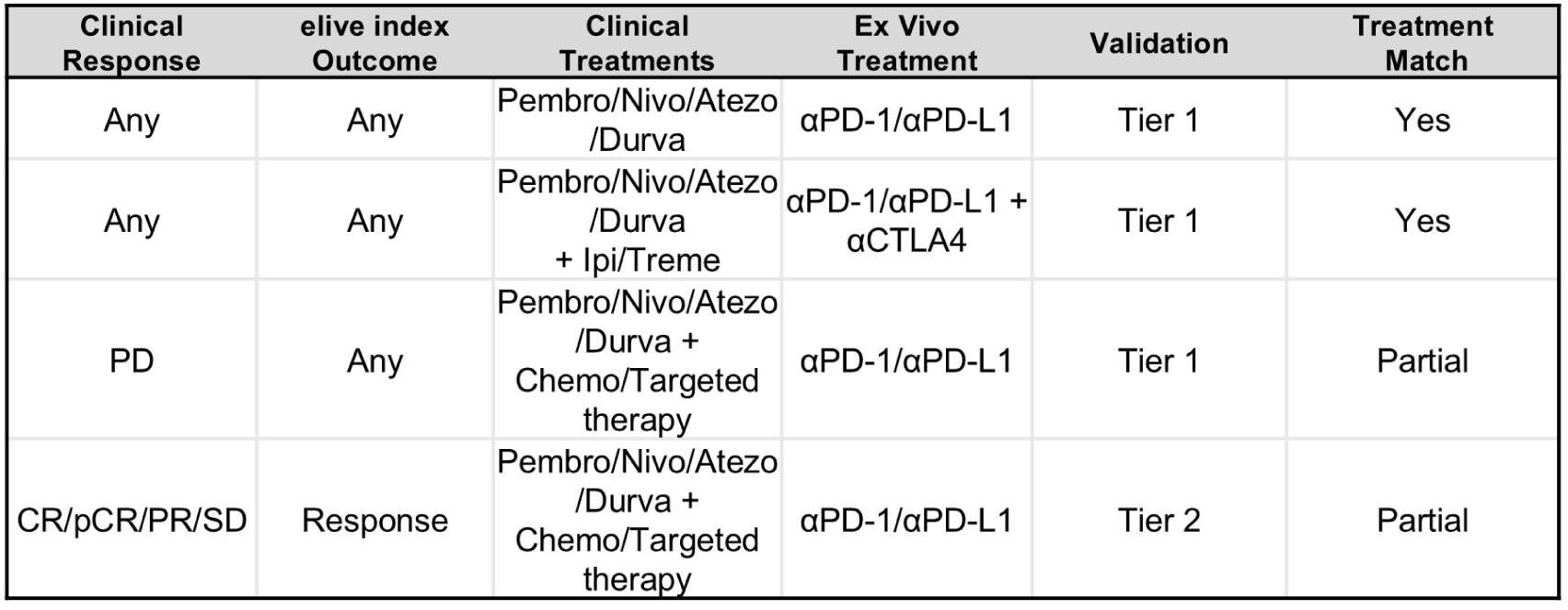
Treatment match and validation calls. Table summarizing conditions of treatment match and validation type criteria.

| Clinical Response | elive index Outcome | Clinical Treatments | Ex Vivo Treatment | Validation | Treatment Match |
| --- | --- | --- | --- | --- | --- |
| Any | Any | Pembro/Nivo/Atezo /Durva | αPD-1/αPD-L1 | Tier 1 | Yes |
| Any | Any | Pembro/Nivo/Atezo /Durva + Ipi/Treme | αPD-1/αPD-L1 + αCTLA4 | Tier 1 | Yes |
| PD | Any | Pembro/Nivo/Atezo /Durva + Chemo/Targeted therapy | αPD-1/αPD-L1 | Tier 1 | Partial |
| CR/pCR/PR/SD | Response | Pembro/Nivo/Atezo /Durva + Chemo/Targeted therapy | αPD-1/αPD-L1 | Tier 2 | Partial |

